# Implementing essential diagnostics-learning from essential medicines: A scoping review

**DOI:** 10.1101/2022.07.04.22277153

**Authors:** Moriasi Nyanchoka, Mercy Mulaku, Bruce Nyagol, Eddy Johnson Owino, Simon Kariuki, Eleanor Ochodo

## Abstract

**Background:** The World Health Organization (WHO) model list of Essential In vitro Diagnostic (EDL) introduced in 2018 complements the established Essential Medicines List (EML) and improves its impact on advancing universal health coverage and better health outcomes. We conducted a scoping review of the literature on the implementation of the WHO essential lists in Africa to inform the implementation of the recently introduced EDL.

**Methods:** We searched eight electronic databases for studies reporting on the implementation of the WHO EDL and EML in Africa. Two authors independently conducted study selection and data extraction, with disagreements resolved through discussion. We used the Supporting the Use of Research Evidence (SURE) framework to extract themes and synthesized findings using thematic content analysis. We used the Mixed Method Appraisal Tool (MMAT) version 2018 to assess the quality of included studies where applicable.

**Results:** We included 172 studies reporting on EDL and EML after screening 3,813 articles titles and abstracts and 1,545 full-text papers. Most (75%, n=129) included studies were purely quantitative in design comprising descriptive cross-sectional designs (60%, n=104), 15% (n=26) were purely qualitative, and 10% (n=17) had mixed-methods approaches. There were no qualitative or randomised experimental studies about the EDL. The main barrier facing the EML and EDL was poorly equipped health facilities - including unavailability or stock-outs of essential in vitro diagnostics and medicines and inadequate infrastructure to enable health service delivery. Financial and non-financial incentives to health facilities and workers were a key enabler to the implementation of the EML however, their impact differed from one context to another. Fifty-six (33%) of the included studies were of high quality.

**Conclusions:** The EDL implementation at the national level can learn from health system interventions to improve the availability and supply of essential medicines such as financial and non-financial incentives in different contexts.

**Plain language summary:** The World Health Organization (WHO) periodically publishes the Model lists of essential medicines (EML) and essential in vitro diagnostics (EDL) to offer guidance to member states. The model lists help countries prioritise the critical health products that should be widely available and affordable throughout health systems. Countries frequently use these model lists to help develop their local lists of essential medicines and diagnostics. The model list of essential diagnostics was introduced in 2018, while the essential medicines were introduced 45 years ago. This work evaluates current evidence on the implementation of the more established model list of essential medicines to inform the development and implementation of the national list of essential in vitro diagnostics in Africa.

We reviewed results from all available studies that looked at the provision of treatment and/or diagnostic services in Africa and assessed the barriers and enablers for their implementation.

We found 172 articles assessing the provision of treatment and diagnostics in Africa. We looked in detail at the barriers and enablers to implementing the model lists of essential medicines and essential in vitro diagnostics. We also assessed the quality of the included research studies. We combined the results of the studies and established that the health system barriers were the most dominant constraints to implementing the model lists.

Our review found the implementation of the established EML, the new EDL was mainly due to poorly equipped health facilities, including limited availability, and stock outs of essential medicines and tests. It is important to consider these constraints when developing and implementing the EDL at various national levels. EDL Implementation at the national level can learn from interventions to improve the availability and supply of essential medicines. Financial and non-financial incentives may be enabling interventions, but their effect varies in different countries and contexts.

## Introduction

Access to diagnostic tests is a key component in achieving Sustainable Development Goals (SDG) 3.8. and Universal Health Coverage (UHC) ^1, 2^. Insufficient access to essential in-vitro diagnostics is a major global health challenge, and nearly half (47%) of the global population have little to no access to diagnostics ^3^. The scale and scope of this challenge contribute to delay in diagnosis and initiation of appropriate treatment compromising health outcomes, especially in Africa ^3–13^.

The World Health Organization (WHO) published the first model list of Essential In Vitro Diagnostics (EDL) in 2018 ^14^ to guide the selection and prioritisation of essential diagnostics according to national needs. It complements the WHO Essential Medicines List (EML) and links medicines with diagnostic tests to advance the UHC ^11, 15^. It paves the way toward improved healthcare delivery and ultimately better patient outcomes by promoting greater equitable access to quality and affordable diagnostics at all levels of the healthcare delivery system ^16^. To ensure its impact, countries will need to develop national lists that suit their national or regional needs, disease burdens, and health system capacities ^2, 5, 17^. To date, three WHO EDL model lists have been published ^14, 18, 19^. The first WHO EDL contained 113 tests and was updated by WHO in 2019 to include nine additional tests for non-infectious diseases. The 2020 list had more other tests, including pandemics such as Covid-19.

The WHO published the first EML about 45 years ago, in 1977 ^20^. This established initiative has been updated biannually since 1977, with the latest 22^nd^ version updated in September 2021 ^21^. Though the adaption of WHO EML to National Essential Medicines Lists (NEMLs) has been broad in Africa, numerous challenges continue to blunt its impact, including persistent inadequate and inequitable access to medicines ^22–25^. Lessons learned in implementing the established WHO EML may shed light on implementation considerations of the WHO EDL and guide the development of practice tools to support the wider adoption of the WHO EDL in Africa.

The objective of this scoping review was to map evidence on the implementation and evaluation of the WHO’s essential lists in African countries to guide the effective implementation of the new WHO EDL.

## Methods

The protocol of this scoping review is available on Open Science Framework ^26^ with deviations from the protocol listed in the S1 Appendix.

We conducted this review according to the Joanna Briggs Institute guidelines for scoping reviews ^27^ and adhered to the Preferred Reporting Items for Systematic Reviews and Meta-Analyses extension for Scoping Reviews (PRISMA-ScR) ^28^ checklist recommended for scoping reviews (S1 Checklist).

### Information sources

A systematic search of the literature was conducted up to May 2021 without date restrictions. We searched several electronic databases: *Ovid MEDLINE, Embase, CINAHL, Web of Science, African Index Medicus, Cochrane Central Register of Controlled Trials, SCOPUS, and Health system evidence for eligible studies.* An example of the search strategies *MEDLINE* can be found in the S2 Appendix. The literature search was complemented by scanning the reference lists of included studies. The references were exported to an EndNote database following the literature search, and the duplicates were removed.

### Eligibility criteria

#### Types of studies

We included EDL studies published in English after introducing the first EDL in 2018. However, given the vast number of studies for the 40-year EML initiative, we included EML studies published in 2010 and after to get a recent representative sample that would inform the implementation of the EDL. If data saturation were not achieved in this sample, we would look to studies published before 2010. We included primary experimental and observational studies and primary qualitative studies. We excluded study protocols, literature reviews, systematic reviews, scoping reviews, book chapters, personal opinions papers, editorials, and conference abstracts with insufficient information. We, however, excluded all conference abstracts and editorials on EML due to the vast number of full-text studies and the high likelihood of data saturation.

We selected eligible studies guided by the Population-Concept-Context (PCC) framework designed by the Joanna Briggs Institute ^29^, commonly used to focus research questions for scoping reviews as detailed below:

#### Population

We included articles reporting on the provision of essential medicines and diagnostic tests (as defined by authors or by the WHO Criteria)^19, 21^ to human populations. We did not limit our review to any disease condition.

#### Concept

We included articles that discussed the implementation of the WHO essential lists, defined in our review as the adoption and adaptation of WHO essential lists by individual WHO member states to address national priority healthcare needs and gaps in the health systems. We also included articles that evaluated interventions used to enhance or enable the implementation or uptake of the essential lists.

#### Context/Settings

We included all studies conducted in all health care settings or levels in Africa. Due to the high likelihood of data saturation, we restricted EML studies to those conducted in single countries. Such studies were likely to give rich data about implementation considerations in one setting or context.

#### Outcomes

Our outcomes of interest were:

- Types of study designs about the implementation of the EDL and EML.
- Themes about barriers and enablers of EDL and EML.

### Study selection

We uploaded references compiled using *Endnote* to Covidence ^30^, a web-based systematic review software platform. We first screened titles and abstracts for potentially eligible articles and then screened full texts of the articles where available. Independent reviewers (MN, BN, EJO, MM) screened all titles, abstracts, and full-text articles in duplicate for eligibility. We resolved disagreements through consensus in consultation with a senior reviewer (EO). Articles that met the inclusion criteria following a full-text review were selected for data extraction.

### Data extraction

We developed a data extraction form using the google form platform. We piloted it with 10% of the included studies by two reviewers to ensure the accuracy of the form and consistency of the extracted content.

The extracted data included general study information (authors and year of publication and the study title); general study characteristics (the study aim, study type, country of study, study setting, facility type, the study population), and information about barriers and enablers to implementing the essential lists (EDL and EML). We extracted data independently in duplicate (MN, BN, EJO, MM) about themes for barriers and enablers to health systems using the Supporting the Use of Research Evidence (SURE) framework (S2 Appendix) ^31^. We resolved disagreements in data extraction through consensus in consultation with a senior reviewer (EO).

### Quality assessment

We assessed the methodological quality of all included studies using the Mixed Method Appraisal Tool (MMAT 2018) ^32^. The tool is grouped into five categories of study designs: qualitative designs, quantitative randomized controlled trials, quantitative non-randomized, quantitative descriptive, and mixed methods. The appraisal led to an overall methodological quality rating varying from unclassified, 0% (no quality), 20% (low quality), 60% (moderate quality), 80 (considerable quality), and 100% (high quality) for each study. Not all eligible studies provided sufficient information to appraise quality using the MMAT. A study was categorised unclassified if it was a report or study that did not provide adequate information for MMAT appraisal. Quality ratings were not used to include or exclude studies but instead used to describe the overall quality of evidence of the included studies.

### Data synthesis

Thematic content analysis was conducted to chart codes and synthesize themes. We summarized the results descriptively and graphically.

## Results

### Search results

Our search yielded 3813 records, nine of which were duplicates Fig 1. Of the remaining 3804 citations screened, 2259 did not meet the inclusion criteria. A further 1373 citations were excluded at full text review, as they did not meet the inclusion criteria for full text review based on year of publication (n=523), not focussing on implementation of WHO essential lists (n=324), ineligible article type (n=265), ineligible context (n=102), multi-country studies about EML (n=67), no full text availability (n=52), duplicates (n=21), non-English publication language (n=16), animal studies (n=3).

### Characteristics of included studies

We included 172 studies. Four (2.3%) of the studies were on the implementation of EDL, eleven (6.4%) focused both on EDL and EML, and 157 (91.3%) focused only on the EML. Details of the included studies can be found in the appendix (S3 Appendix).

Of the (EDL) studies (n=15), eight (53.3%) were from Eastern Africa, five (33.3%) from Southern Africa, and two (13.3%) from West Africa. Methodologically, twelve (80%) of EDL studies used quantitative methods (cross-sectional designs), and three (20%) used the mixed methods approach (Table 1).

**Table 1:**
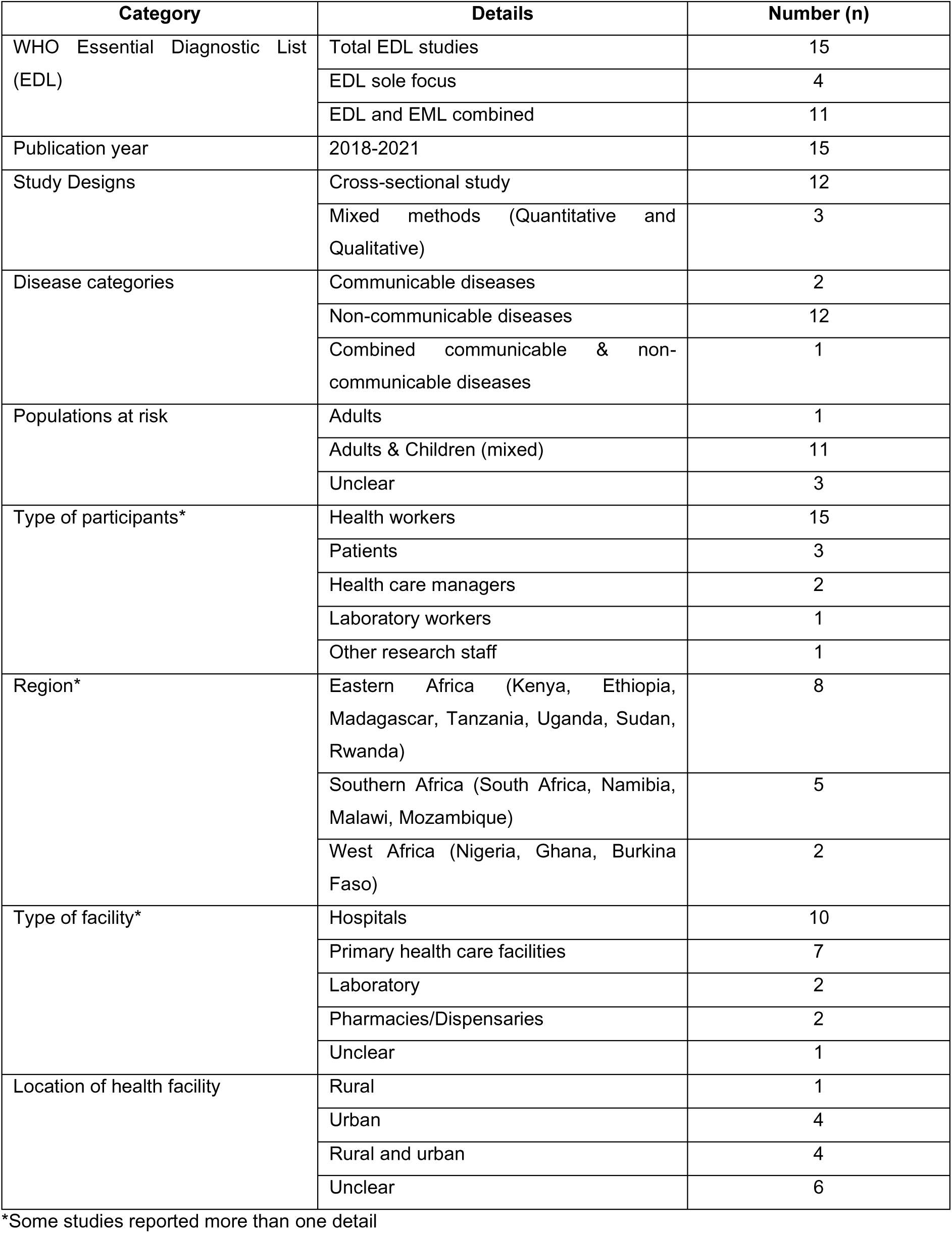
Characteristics of Studies about the Essential In Vitro Diagnostics List (EDL)

| Category | Details | Number (n) |
| --- | --- | --- |
| WHO Essential Diagnostic List (EDL) | Total EDL studies | 15 |
|  | EDL sole focus | 4 |
|  | EDL and EML combined | 11 |
| Publication year | 2018-2021 | 15 |
| Study Designs | Cross-sectional study | 12 |
|  | Mixed methods (Quantitative and Qualitative) | 3 |
| Disease categories | Communicable diseases | 2 |
|  | Non-communicable diseases | 12 |
|  | Combined communicable & non-communicable diseases | 1 |
| Populations at risk | Adults | 1 |
|  | Adults & Children (mixed) | 11 |
|  | Unclear | 3 |
| Type of participants* | Health workers | 15 |
|  | Patients | 3 |
|  | Health care managers | 2 |
|  | Laboratory workers | 1 |
|  | Other research staff | 1 |
| Region* | Eastern Africa (Kenya, Ethiopia, Madagascar, Tanzania, Uganda, Sudan, Rwanda) | 8 |
|  | Southern Africa (South Africa, Namibia, Malawi, Mozambique) | 5 |
|  | West Africa (Nigeria, Ghana, Burkina Faso) | 2 |
| Type of facility* | Hospitals | 10 |
|  | Primary health care facilities | 7 |
|  | Laboratory | 2 |
|  | Pharmacies/Dispensaries | 2 |
|  | Unclear | 1 |
| Location of health facility | Rural | 1 |
|  | Urban | 4 |
|  | Rural and urban | 4 |
|  | Unclear | 6 |

\*Some studies reported more than one detail

Of the EML studies (n=168), eighty-five (51.2%) were from Eastern Africa, forty-five (27.4%) were from Southern Africa, thirty-two (19.0%) were from West Africa, five (3.0%) from Central Africa and one (0.6%) from North Africa (Table 2).

**Table 2:** Characteristics of studies about the Essential Medicine List (EML)

| Category | Details | Number (n) |
| --- | --- | --- |
| Essential Medicine List (EML) | Total EML studies | 168 |
|  | EML focus only | 157 |
|  | Combined EML & EDL studies | 11 |
| Publication year | 2010 – 2021 | 168 |
| Study Designs | Cross-sectional study | 100 |
|  | Qualitative study | 26 |
|  | Mixed methods | 17 |
|  | Cohort study | 5 |
|  | Randomized controlled trial | 4 |
|  | Interrupted Time Series | 4 |
|  | Quasi-Experimental study | 1 |
|  | Other Experimental designs | 2 |
|  | Other Descriptive designs | 9 |
| Disease categories | Communicable diseases | 19 |
|  | Non-communicable diseases | 51 |
|  | Both communicable and non-communicable diseases | 51 |
|  | Unclear | 47 |
| Populations at risk | Children | 22 |
|  | Adults | 16 |
|  | Mixed populations (adults and children) | 86 |
|  | Unclear | 44 |
| Type of participants* | Patients | 33 |
|  | Health workers | 129 |
|  | Health care managers | 31 |
|  | Laboratory workers | 2 |
|  | Community residents | 8 |
|  | National Essential Lists Committee | 15 |
|  | Health care stakeholders | 13 |
|  | Policy document analysis, reviews, Inventory analysis, Ministry officials | 15 |
|  | Unclear | 7 |
| Region | North Africa | Egypt (1) |
|  | West Africa | Benin, Burkina Faso, Ghana, Mali, Nigeria, Sierra Leone (32) |
|  | Central Africa | Cameroon, Gabon (5) |
|  | Eastern Africa | Ethiopia, Kenya, Madagascar, Rwanda, Sudan, Tanzania, Uganda (85) |
|  | Southern Africa | Botswana, Eswatini, Malawi, Mozambique, Namibia, South Africa, Zambia, Zimbabwe (45) |
| Type of facility* | Hospitals | 95 |
|  | Primary Health care facilities | 90 |
|  | Pharmacies/Dispensaries | 45 |
|  | Community services | 5 |
|  | Others (Clinics, Medical centres, National essential list committees) | 4 |
|  | Unclear | 13 |
| Location of health facility* | Rural | 25 |
|  | Peri-Urban | 11 |
|  | Urban | 26 |
|  | Rural and Urban | 36 |
|  | Peri-urban and Urban | 6 |
|  | Rural, Urban, Peri-Urban | 4 |
|  | Unclear | 74 |

\*Some studies reported more than one detail

### Study designs of included studies

Overall, most of the studies (n=129, 75%) in our review used quantitative study designs, followed by qualitative (n=26, 15%) and mixed methods (n=17, 10%). Studies with quantitative methods were mainly descriptive cross-sectional designs (n=104, 60%), followed by experimental or intervention studies (n=11, 6%) and cohort study designs (n=5, 3%). A summary of EDL study designs is presented in Table 1, and EML studies in Table 2.

All studies with a sole focus on EDL were cross-sectional study designs. Most studies on EML were quantitative studies (n=125, 73%) and mostly descriptive cross-sectional studies.

### Themes about barriers and enablers

Barriers and enablers facing the EDL and EML were similar and were mostly about health system constraints. We present key themes about the barriers and enablers of the EML and EDL stratified in SURE themes below: At the individual level, themes about recipients of care, providers of care, and other stakeholders included their knowledge and skills, attitudes, and motivation to implement the WHO essential lists. Themes about health systems were myriad and will focus on the results section. Themes about social and political levels included ideologies, contracts, legislation or regulations, donor policies, influential people, corruption, and political stability.

### Barriers and enablers for the implementation of the EDL and EML

A summary of themes about barriers facing the EDL and EML in the health system domain is presented in Fig 2. Themes about health systems constraints facing the EDL and EML were similar in included studies and mainly were about health system-level constraints. They included: limited availability of essential medicines and diagnostics at primary health care facilities compared to higher-level health facilities, lack of human resources, limited access to care facilities by patients due to financial and geographical constraints, procurement and distribution systems leading to regular stockouts, poorly resourced health facilities due to limited health facility operational funds, inefficient information systems and limited staff training. These barriers challenge implementing essential lists, especially the novel WHO EDL across countries. Other significant challenges unique to the EDL included lack of proper equipment and supplies, inadequate infrastructure, and space to facilitate laboratory and diagnostics services and a shortage of skilled laboratory staff to support the implementation of the EDL at different health care levels as guided in the WHO EDL. The prominent themes about barriers to the implementation of the EDL and EML are presented in Table 3 and Table 4, respectively.

**Table 3:** Thematic analysis of the 9 applied SURE codes to barriers for the implementation of EDL

| Proportion of total themes per code |  |
| --- | --- |
| **Health system constraints** |  |
| Facilities (N=9, n=17) |  |
| Unavailability of tests | 11 (65%) |
| Reagent stock-outs | 3 (18%) |
| Lack of proper equipment | 2 (12%) |
| Inadequate infrastructure and space | 1 (6%) |
| Accessibility of care (N=6, n=8) |  |
| Poor financial access | 4 (50%) |
| Poor health facility access | 2 (25%) |
| Poor geographical access | 1 (12.5%) |
| Poorly resourced health facilities | 1 (12.5%) |
| Procurement and distribution (N=3, n=3) |  |
| Poor supply chain management including poor quantification and low inventory levels | 3 (100%) |
| Human resources (N=3, n=3) |  |
| Shortage of laboratory staff | 1 (33%) |
| Inadequate number of qualified and skilled lab staff | 1 (33%) |
| General shortage of health worker | 1 (33%) |
| Information systems (N=3, n=3) |  |
| Absence of clinical case registries | 2 (67%) |
| Unavailability of operational data | 1 (33%) |
| Financial resources (N=2, n=2) |  |
| Inadequate health facility operational funds | 1 (50%) |
| High test costs | 1 (50%) |
| Education system (N=2, n=2) |  |
| Improper training of laboratory staff | 1 (50%) |
| Lack of training | 1 (50%) |
| Relationship with norms and standards (N=1, n=1) |  |
| Poor availability of guidelines | 1 (100%) |
| **Social and Political constraints** |  |
| Legislation or regulations (N=1, n=1) |  |
| Insufficient policy | 1 (100%) |

N=number of studies that cited the SURE theme, n= frequency of applied codes per SURE theme SURE= Supporting the Use of Research Evidence

**Table 4:** Thematic analysis of the most applied SURE codes to barriers for the implementation of EML

| Proportion of total themes per code |  |
| --- | --- |
| **Health system constraints** |  |
| Facilities (N=76, n=81) |  |
| Low availability of essential medicines in health facilities | 45 (56%) |
| Unavailability of essential medicines in public and lower-level health facilities | 15 (19%) |
| Inadequate health facility capacity | 10 (12%) |
| Human resources (N=44, n=62) |  |
| Lack of human resource | 19 (31%) |
| Inadequate capacity of health workers | 11 (18%) |
| Low number of specialized healthcare workers | 8 (13%) |
| Accessibility of care (N=35, n=53) |  |
| Poor financial access | 22 (42%) |
| Poor geographical access | 16 (30%) |
| Poor health facility access | 15 (28%) |
| Procurement and distribution systems (N=40, n= 49) |  |
| Inefficient procurement processes | 16 (33%) |
| Poor stock management | 10 (20%) |
| Inefficient distribution systems | 9 (18%) |
| Relationship with norms and standards (N=33, n= 35) |  |
| Unavailability of guidelines | 16 (46%) |
| Incompliance to guidelines | 8 (23%) |
| Incompliance to national EMLs | 6 (17%) |
| Financial resources (N=25, n=25) |  |
| Insufficient health facility funding | 22 (88%) |
| Late claimant rebates from national insurance agencies | 2 (8%) |
| Poor effect of financial autonomy | 1 (4%) |
| Information systems (N=18, n=18) |  |
| Poor information management practices | 7 (39%) |
| Lack of reporting procedures and ordering systems | 4 (22%) |
| Inadequate information systems | 4 (22%) |
| Clinical supervision (N=11, n= 11) |  |
| Lack of supportive supervision | 9 (82%) |
| Inadequate health worker supervision | 2 (18%) |
| Educational system (N=10, n=10) |  |
| Lack of staff training | 6 (60%) |
| Lack of specialized training | 3 (30%) |
| Lack of educators to train health workforce | 1 (10%) |
| Allocation of authority (N=10, n= 10) |  |
| Limited facility manager authority on facility budget, procurement, pricing, and supply of medicines | 7 (70%) |
| Lack of health workers involvement in drug selection and procurement | 2 (20%) |
| Lack of autonomy by community members and facility managers on the community health fund | 1 (10%) |
| Accountability (N=8, n=10) |  |
| Lack of accountability of authorities on medicine orders, procurement, distribution, and stock management | 10 (100%) |
| Management and or leadership (N=9, n=9) |  |
| Inadequate leadership and coordination capacity | 7 (78%) |
| Inadequate leadership support to rural-based health workers | 1 (11%) |
| Lack of knowledge and skills by health workers working in managerial roles | 1 (11%) |
| Internal communication (N=9, n=9) |  |
| Lack of coordination among stakeholders, health managers, and health facilities | 6 (67%) |
| Poor communication of policy change | 2 (22%) |
| Limited communication between health facilities | 1 (11%) |
| **Incentives (N=6, n=6)** |  |
| Low motivation of health workers including high workload, poor remuneration, and non-payment of stipends | 4 (67%) |
| Non-payment of suppliers | 1 (16%) |
| Low tax incentives and subsidies to pharmaceuticals | 1 (16%) |
| **Bureaucracy (N=5, n=5)** |  |
| Bureaucratic decision making with limited evidence | 5 (100%) |
| **Patient flow processes (N=5, n=5)** |  |
| Poor referral practices | 5 (100%) |
| **External communication (N=2, n=2)** |  |
| Lack of information/communication material for provision of standard care | 1 (50%) |
| Insufficient levels of essential information for consumers | 1 (50%) |
| **Social and Political constraints** |  |
| **Legislation or regulations (N=34, n=36)** |  |
| Lack of pricing policy | 10 (28%) |
| Incompliance to regulations | 7 (19%) |
| Lack of structured guidelines | 5 (14%) |
| **Donor policies (N=6, n=6)** |  |
| Donors influence on the implementation of EML | 4 (67%) |
| International procurement policies of donors | 1 (17%) |
| Poor policy adoption from the global to national level | 1 (17%) |
| **Influential people (N=5, n=6)** |  |
| Pharmaceutical promotions to influence prescription practice and revisions of standard treatment guidelines | 2 (33%) |
| International recommendations | 1 (17%) |
| Lack of support to local pharmaceutical production | 1 (17%) |
| **Ideology (N=4, n=4)** |  |
| Market ideologies | 1 (25%) |
| Political ideologies | 1 (25%) |
| Community beliefs and attitudes | 1 (25%) |
| **Corruption (N=4, n=4)** |  |
| Corruption practices in public companies | 3 (75%) |
| Trading of counterfeit medicines | 1 (25%) |
| **Contracts (N=3, n=3)** |  |
| Absence of national medical contracts | 1 (33%) |
| Delay in contracting pharmaceutical tenders | 1 (33%) |
| Absence of structure contracts | 1 (33%) |
| **Political stability (N=1, n=1)** |  |
| Political instability | 1 (100%) |
| **Individual level constraints** |  |
| **Providers of care** |  |
| **Knowledge and skills (N=21, n = 21)** |  |
| Inadequate training on current evidence-based treatment | 8 (38%) |
| Insufficient number of skilled healthcare workers | 5 (24%) |
| Inadequate providers knowledge on disease management | 3 (14%) |
| **Motivation to change or adopt new behaviour (N=8, n=10)** |  |
| Underpayment of healthcare workers | 3 (30%) |
| High workload | 2 (20%) |
| Limited supervision, career, and training opportunities | 2 (20%) |
| **Attitudes towards programme (N=6, n= 6)** |  |
| Quality concerns of medicines from some manufacturers | 1(17%) |
| Negative attitude towards new quality improvement projects | 1(17%) |
| Poor perceptions and awareness of child-appropriate medicine dosage formulations | 1(17%) |
| **Recipients of care** |  |
| **Attitudes towards programme (N=9, n=10)** |  |
| Preference for private pharmacies to primary health care centres | 2 (20%) |
| Social cultural influences | 2 (20%) |
| Use of alternative treatments | 2 (20%) |
| **Motivation to change or adopt new behaviour (N=6, n=7)** |  |
| Unaffordability of medicines | 2 (29%) |
| Low availability of drugs in health facilities | 1 (14%) |
| Poor access to health facilities | 1 (14%) |
| Knowledge and skills (N=2, n=2) |  |
| Inadequate consumer knowledge on drugs | 1 (50%) |
| Insufficient understanding of diseases | 1 (50%) |
| **Other stakeholders** |  |
| Knowledge and skills (N=5, n= 5) |  |
| Inadequate training on rational use of medicines | 2 (40%) |
| Inadequate knowledge on health commodities and financial report | 1 (20%) |
| Unequal access to information on medical products | 1 (20%) |
| Motivation to change or adopt new behaviour (N= 4, n=4) |  |
| Inadequate drugs and medical supplies | 2 (50%) |
| Low motivation to participate in healthcare programmes | 1 (25%) |
| Poor health worker and patient relationships | 1 (25%) |

N=number of studies that cited the SURE theme, n= frequency of applied codes per SURE theme SURE= Supporting the Use of Research Evidence

The themes about enablers for implementing both the EML and EDL are opposite to the health system barriers listed above. They are about tackling the listed barriers and have been summarised in Fig 3.

### Individual-level

In this section, we present individual-related themes identified for the implementation of the EML.

#### Providers of care

##### Knowledge and skills

Barriers related to knowledge and skills were the most prominent theme under the providers of care domain. The main barriers identified included inadequate training on current evidence-based treatment ^33–40^, an insufficient number of skilled healthcare workers ^41–45^, and inadequate providers’ knowledge of disease management ^46–48^. Other barriers included lack of knowledge of inventory management ^49, 50^, lack of awareness of available guidelines ^51^, and poor understanding of partner programmes^52, 53^.

The knowledge and skills-related enablers for the implementation of the EML are opposite to the main barriers listed above ^54–59^.

##### Motivation to change

The main barriers to motivation to change included underpayment of healthcare workers ^60–62^, high workload as a result of shortage of staff ^63, 64^ and limited supervision, career and training opportunities ^62, 65^. Other motivation related barriers cited include low levels of motivation to work due to lack of essential medicines and equipment for use in facilities ^36^, delay in the payment of claimant rebates to health staff ^61^, and limited scientific evidence-informed decision-making ^66^.

The provision of financial incentives to staffs in the pay for performance (P4P) programmes was identified as an enabler for the implementation of the EML ^67^.

##### Attitudes regarding programme acceptability, appropriateness, and credibility

The identified barriers related to the attitudes of providers of care towards implementation of EML included quality concerns of medicines from some manufacturers ^68^, negative staff attitudes towards new quality improvement projects ^36^, poor perceptions and awareness of child-appropriate medicine dosage formulations ^46^, limited prescription of pain medication and stigmatization of palliative care ^45^, consideration of traditional medicines on other illnesses ^69^, and concerns on adopted ICT systems for inventory management ^70^.

The enablers identified related to the providers of care attitudes towards the implementation of the EML included satisfaction with the coverage of the social health insurance scheme ^61^, positive attitude towards the provision of ICT support to improve their medical knowledge and skills ^71^, and satisfaction with the quality of medicines ^72^.

#### Recipient of care

##### Attitudes regarding programme acceptability, appropriateness, and credibility

Patients’ attitudes regarding acceptability, appropriateness and credibility related to EML implementation were the most prominent theme under the recipient of care. The main barriers reported include preference to seek care from private pharmacies than primary health care centres due to lack of essential drugs and beliefs on the quality of medicines ^68, 73^, social-cultural influences ^74, 75^, and use of alternative treatments (traditional medicine) ^68, 73^. Other barriers reported include perception of the unaffordability of drugs ^76^, uncertainty on availability of services ^77^, drug safety concerns ^64^, and low health-seeking behaviour ^76^.

##### Motivation to change

The reported barriers related to motivation to change were unaffordability of medicines ^78, 79^, low availability of drugs in health facilities ^76^, poor accessibility to the facility ^74^, poor patient and health workers relations ^41^, lack of social and welfare support ^74^, and inadequate supplies in health facilities ^80^. The motivation to change related enablers for the implementation of the EML are opposite to the barriers reported above ^34, 52, 58, 75, 76, 81–86^.

##### Knowledge and skills

The barriers to receiving care knowledge and skills related to the EML implementation were mainly inadequate consumer knowledge of medical and technical information on drugs they purchase and insufficient understanding of disease management ^87, 88^.

The recipients of care knowledge and skills-related enablers for the implementation of the EML included adequate on prescribed drugs ^59^ and adequate knowledge on conditions ^34, 89^.

**Other stakeholders** (community health committees, community leaders, programme managers, donors, policymakers, opinion leaders)

##### Knowledge and skills

The barriers related to other stakeholders’ knowledge and skills reported include inadequate training on rational use of medicines ^90, 91^, inadequate knowledge of health commodities and financial reports amongst health facility and governing committee members ^92^, unequal access to information on medical products to all stakeholders ^88^, and variation in the knowledge of child-appropriate dosage formulations among stakeholders ^46^.

Knowledge and skills on the quality, safety, and efficacy of medicines and pharmacoeconomic evaluations by stakeholders in selecting medicines for the EML were cited as an enabler for the implementation of the EML ^93^.

##### Motivation to change

Barriers related to the motivation to change are mainly inadequate drugs and medical supplies, limiting the provision of quality care ^75, 94^, low motivation to participate in healthcare programmes ^95^, and poor health worker and patient relationships leading to poor health-seeking behaviour ^82^.

On the other hand, the provision of incentives to stakeholders was identified as an enabler of implementing EML. These included the provision of business incentives to owners of accredited drug dispensing outlets (ADDOS) ^55^ and financial incentives through payment for performance (P4P) to health facilities, district and regional managers ^67^ to improve service delivery, availability of medicines and medical supplies in poor and rural areas.

### Health systems-level

#### Facilities

The most reported barrier to implementing WHO essential lists (EDL&EML) was the facility-related constraints (Table 3 and Table 4). Unavailability of EDL tests ^6, 96–102^, and reagent stock-outs ^6, 100, 103^ were the most prominent themes within the facility-related barriers to EDL implementation. Other EDL barriers referenced lack of proper equipment and supplies described as low availability of key consumables for laboratory diagnosis, limited items of the major laboratory equipment’s ^6, 96^, and inadequate infrastructure and space ^96^ to facilitate laboratory and diagnostics services. Similarly, in the EML implementation, the most prominent themes within this barrier were low availability and unavailability of essential medicines ^38, 40, 46, 51, 82, 89, 104–112^.

The enabler themes for the EDL implementation are opposite to the EDL barriers mentioned above ^103, 113–115^. The EML enablers are opposite of the EML barriers reported above, and mainly included availability of essential medicines in facilities^34, 37, 48, 57–59, 66, 87, 92, 108, 116–137^, and adequate capacity of facilities to provide care ^34, 138^.

#### Accessibility of care

Accessibility of care-related barriers to EDL implementation was one of the most prominent barriers identified at the health system-level. The most prominent theme under this EDL barrier was poor financial access. This included limited access to care facilities by patients due to financial constraints: out of pocket expenses ^103, 139^ and high test costs ^96, 99^. Other EDL barriers were poor geographical access contributing to limited access to essential in vitro diagnostics in rural areas ^96^, and limited health facility access: availability of essential in vitro diagnostics at referral hospitals but not primary health care facilities ^96^ and public facilities ^98^, and poorly resourced health facilities ^97^. The barriers for the implementation of the EML are similar to the EDL barriers reported above, and the main barriers were poor financial access ^39, 48, 78, 79, 83, 86–89, 104, 110, 136, 140–149^, poor geographical access ^36, 65, 74, 82, 104, 143, 146, 150^ and health facility access ^73, 74, 106, 150–152^.

The accessibility of care related-enablers for the implementation of both the EDL ^96, 98, 103^ and EML ^48, 50, 53, 57, 64, 68, 69, 79, 83–85, 87, 104, 128, 133, 134^, are opposite to the barriers listed above.

#### Human resources

The human resources-related barriers were a prominent barrier to implementing the WHO essential lists. Shortage of laboratory staff ^96^, inadequate number of qualified and skilled laboratory staff ^114^, and a general shortage of health care workers ^100^ were identified as the main barriers to implementing EDL. Similar human resources-related barriers to the EDL were reported in the implementation of the EML. Barriers unique to the EML included limited staff training ^34, 35, 47, 51, 105, 107, 131, 153^, high workload ^63, 75, 111, 123, 143, 154^ and low number of specialized health care workers ^106, 116, 125, 154^. Other cited EML barriers included inequitable access to health care workers in rural areas ^37, 43, 57^, inequitable training in lower-level facilities ^119^, and inadequate capacity of the essential medicines committee ^66^.

Human resources-related enablers for the implementation of EML reported include sufficient capacity of health care workers ^138, 155^, adequate training provided to health care workers ^58, 156^, and availability of health care workers ^57^.

#### Procurement and distribution systems

Poor supply chain management including poor quantification ^6, 102^ and low inventory levels ^101^ leading to stock-out supplies in health facilities were identified as barriers to EDL implementation. Similar and other related barriers were also cited in the EML implementation. The main barriers unique to the EML included inefficient procurement processes ^34, 47, 49, 63, 65, 69, 70, 82, 157^, poor stock management practices^39, 49, 50, 53, 94, 105, 107, 140, 158, 159^, and inefficient distribution systems ^40, 49, 75, 94, 111, 131, 146, 160, 161^.

Other barriers highlighted included poor quantification of medicines and medical supplies at facility level ^49, 87, 131, 160, 162^, and kits from national medical stores ^115, 163^, limited procurement funding for essential medicines ^46, 93^, lack of training of staffs in procurement ^146^, distribution monopoly by national medical stores ^71^, inappropriate selection of medicines ^131^, and poor monitoring and evaluation ^49^.

Adequate inventory management was identified as an enabler in the EDL implementation ^101^. The EML enablers were identified are the opposite of the EML barriers highlighted above ^49, 50, 62, 64, 68, 70, 92, 121, 122, 130, 164^.

#### Relationship with norms and standards

The barriers related to the relationship with norms and standards for the EDL implementation included poor availability of diagnosis and management guidelines, noting the unavailability of diabetes guidelines in any surveyed clinics ^165^. A similar barrier was identified in the implementation of the EML. Unavailability of guidelines was the most prominent theme within the barriers related to relationship with norms and standards for the EML implementation ^35, 59, 66, 86, 106, 110, 116, 119, 131, 143, 154, 155, 166, 167^. Other barriers unique to the EML included incompliance to guidelines ^50, 53, 131, 149, 153, 166, 168, 169^, incompliance to national EMLs ^48, 68, 79, 132, 145, 170^, lack of national EMLs in facilities ^128, 171^, disconnect between guidelines on treatment protocols ^47, 75^, disconnect between policy, guidelines and practice ^46, 167^, and lack of standard operating procedures ^135^.

The enablers for the EDL and EML are opposite to the barriers mentioned above ^33, 63, 68, 79, 92, 113^. The enabler unique to the EML was compliance with the national EML in rural settings compared to urban ^68^.

#### Financial resources

Inadequate health facility operational funds ^96^ and high test costs due to much greater resource requirements depending on the type of diagnostics test ^99^ were identified as barriers to EDL implementation. Similar financial constraints were also reported in implementing the more established EML. Inadequate funding was the most prominent theme within financial constraints barrier to the implementation of EML^34, 43, 46, 94, 95, 109, 110, 131, 137, 146, 149, 153, 154, 158, 159, 163, 172–175^.

The financial resources-related enablers for the implementation of the EML are opposite to the barriers reported above ^120, 121, 123^.

#### Information systems

The information systems-related barriers to EDL implementation identified include the absence of clinical case registries ^97, 115^ and the unavailability of operational data on EDL accessibility in some facilities ^101^. Similar barriers were identified in the implementation of EML ^43, 49, 65, 173^. Other barriers unique to the EML included poor information management practices ^50, 110, 111, 122, 125, 137, 142^ consisting of poor record-keeping, lack of standardized treatment protocols in health facilities, lack of reporting procedures and structured ordering systems ^107, 131, 146, 176^, and inadequate capacity to support information systems ^70, 105, 138^.

The identified EML enablers were the opposite of the EML barriers. They mainly included good information practices ^55, 67, 177, 178^, access to record-keeping tools ^34^, and utilization of information and communications technology (ICT) ^68, 129^ to monitor real-time data on health facility drug consumption and stock levels.

#### Education system

The education system-related barriers to the essential list’s implementation were related to the training of health workers. The EDL implementation barriers reported include improper staff training to provide laboratory tests ^96^ and health workers’ knowledge gaps, and lack of training ^97^. Similar education system-related barriers are reported in EML studies ^35, 49, 66, 106, 116, 125, 134, 138, 179^. Other barriers unique to the EML included lack of specialized training ^116, 125, 138^, lack of training in the use of economic evidence essential in the selection of medicines ^66^, and lack of educators to train the health workforce ^134^.

Some studies identified education system-related enablers for the implementation of the EML. These included improved access to training ^36, 63, 92^ and in-service training to health workers ^71, 77^.

#### Clinical supervision

The main clinical supervision-related barriers to the EML implementation cited include lack of supportive supervision ^51, 63, 68, 73, 77, 111, 153, 156, 159^ and inadequate health worker supervision ^65, 130^ to support the implementation of EML.

On the other, other EML studies cited regular supportive supervision as an enabler in implementing the EML ^34, 58, 59, 92^.

#### Allocation of authority

The allocation of authority-related barriers was only identified in EML and was related to decision making autonomy to support the implementation of EML. The main barrier identified includes limited authority by the facility managers on facility budget, procurement, pricing, and supply of medicines ^70, 87, 95, 110, 160, 179, 180^. Other constraints included a lack of health workers’ involvement in drug selection and procurement at facility level ^50, 149^ and a lack of autonomy by the community members and facility managers on the community health fund ^94^ to support health care programmes.

Allocation of authority was also reported as an enabler for EML implementation. Direct health financing to a facility led to a successful implementation of a prime vendor system (PVS) due to financial autonomy and flexibility in using funds ^92^.

#### Accountability

Lack of accountability was highlighted as a barrier to EML implementation. These included a lack of accountability of authorities on medicines orders, procurement, distribution, stock management, and delay in disbursement of health funds ^94, 109, 110, 142, 156^. Poor accountability of personnel in the ministry of health and community-based health insurance financing ^87, 110, 156, 164^ and inconsistent inventory records ^111^ in the facility were also highlighted as barriers to EML implementation.

Some studies cited sufficiently structured systems ^92^ and an adequate tracking/management system ^84^ were enablers of accountability in the health system and implementation of EML.

#### Management and leadership

Management or leadership in the health systems was reported as a barrier to EML implementation. Reported barriers include inadequate leadership and coordination capacity to coordinate the support and collaboration by partners and stakeholders ^35, 46, 63, 77, 92, 179, 181^, inadequate leadership support to rural-based health workers ^153^ and lack of knowledge and skills among health workers working in managerial roles ^36^ to further the implementation of EML.

Strong leadership and commitment of the ministry of health ^121^ and facility leadership were identified as enablers for implementing EML ^72^.

#### Internal communication

Lack of coordination among stakeholders, health managers, and health facilities was identified as the primary internal communication-related barrier to implementing EML ^65, 70, 71, 77, 146, 181^. Other identified EML barriers included poor communication of policy change leading to inconsistencies between the procurement list and national essential lists ^50, 166^ and limited communication between health facilities ^41^.

Internal communication-related enablers to the EML included sufficient coordination and communication between health facilities and national medicine stores, health managers, and health care workers ^129, 160^.

#### Incentives

The low motivation of health care workers was the main incentives-related barrier to the EML implementation. This was related to high workload, poor remuneration, and non-payment of stipends ^36, 41, 75, 158^. Other barriers reported include non-payment of suppliers ^70^ and low tax incentives and subsidies to pharmaceutical companies ^170^ to support EML implementation.

The EML enablers identified included providing financial incentives to health system actors (facilities, health managers, and health care workers) through incentivised programmes and interventions that positively affected the quality of health service. These programmes included pay for performance (P4P) and performance-based financing (PBF), innovative financing strategies that provide financial incentives to health service entities and healthcare providers to achieve increased coverage of quality health services. These programmes, P4P ^67, 182^ and PBF ^62, 183, 184^, contributed to improved accessibility of care, availability of essential medicines in health facilities, motivated health care workers and managers, improved health information systems, and greater financial autonomy for health facilities, and increased accountability within the health system.

#### Bureaucracy

Unsupportive bureaucracy was identified as a barrier to the EML implementation. Studies reported bureaucratic decision-making with little or no evidence regarding procurement and supply of essential medicines ^46, 87, 94, 105, 166^.

#### Patient flow processes

The patient flow processes-related barriers to the EML were related to inadequate referral systems and poor referral practices ^41, 75, 77, 112, 146^. These included patients’ referrals conducted without receiving pre-referral treatment and without referral letters affecting the provision of care and treatment ^41, 75, 77, 112, 146^.

An adequate referral system was cited as an enabler to EML implementation ^152^.

#### External communication

Identified EML barriers were related to poor communication practices between health workers and recipients of care. These included lack of information/education/communication material for provision of standard care ^143^ and insufficient levels of essential information for consumers when buying from drug shops^88^.

External communication was identified as an enabler for the implementation of EML are opposite to the EML barriers mentioned above ^34, 36, 83, 85, 112, 128, 156, 179^.

### Social and Political-level

#### Legislation or regulations

The most frequently reported barrier at the social and political level was related to legislation or regulations (Table 3 and Table 4). Insufficient policy to facilitate access to essential diagnostics was identified as a barrier to the EDL implementation ^97^. Barriers unique to the EML included lack of price regulations or pricing policy ^78, 86, 89, 108–110, 141, 148, 174, 180^, incompliance to regulations ^38, 46, 88, 133, 145, 176, 185^, lack of structured guidelines for registration and control ^72, 73, 116, 126, 186^. Other barriers included lack of policies ^73, 153, 155^, inadequate policies that provide control and use of medicines ^44, 53, 187^, long registration process ^68, 146, 170^, restriction on the use of medicines ^41, 142^, lack of political will in implementing policies ^159^, lack of a regulatory body for certifying and professionalizing medical, logistical companies ^159^, and inadequate procedures ^149^.

The EML enablers reported include supportive health financing policy reforms ^142, 188^ and the presence of a structured registration process ^66^ that supported the implementation of the EML.

#### Donor policies

Donor policies were also cited as barriers to the EML implementation. The donors’ influence on EML implementation was reported as a primary barrier. This type of barrier referenced provision of donations irrespective of need, lack of donor priority and operation of donor-funded goods outside registration frameworks ^50, 128, 146, 164^. Other barriers referenced international procurement policies of donors and NGOs disadvantage local producers ^170^, and poor policy adoption from the global to national level ^46^ as barriers to EML implementation.

Donor’s collaboration with government and faith-based organizations was highlighted as an enabler that improved the availability of essential malarial medicines in health facilities ^133^.

#### Influential people

The influential people-related barriers to the EML implementation included pharmaceutical promotions to influence prescription practice and revisions of standard treatment guidelines ^50, 66^ and implementation of international recommendations separate from other national programmes, for instance, the vertical programmes ^66^. Other barriers included the limited influence of health care managers on policy and resource development ^36^, lack of support to the local pharmaceutical production leading to reliance on importations ^68^, and donors’ operation outside the policy framework ^46^.

On the other hand, the influence of government agencies on the wholesale-to-retail market ^88^ and international organizations described as WHO technical support to the government on the guidance and development of WHO model lists, and WHO classification of antimicrobial ^175^ were identified as enablers to the implementation of the EML.

#### Ideology

Barriers to the EML implementation included market ideologies where generic medicines enter the market without a review process ^142^, and political ideologies where facilities are built for political and economic purposes and without the involvement of the communities ^73^ and a disconnect between insurance drug list and drug prescription guidelines ^41^. Community beliefs and attitudes negatively influenced the use of internationally controlled essential medicines (ICEMs) by patients with end-stage diseases ^146^.

#### Corruption

The main corruption-related barrier to the implementation of EML referenced was corruption practices in public companies. This is coupled with awarding tenders to companies without the capacity to supply medical supplies, providing financial incentives to pharmacists from pharmaceutical to dispense specific drugs, and tenders ^68, 70, 72^. The trading of counterfeit medicines was also identified as a barrier to implementing EML ^124^.

#### Contract

The contract-related barriers were only identified in EML. The barriers identified include the absence of national medical contracts in regional code list ^160^, delay in contracting pharmaceutical tenders ^160^, and absence of structured contracts leading to awarding tenders to companies without the capacity to deliver ^70^.

#### Political stability

Political unrest was identified as a barrier to the implementation of the EML. It impacted health care workers staffing in the affected areas ^36^.

### Quality of evidence

One hundred and sixty-seven (97%) articles were appraised for quality. In this scoping review, 56 (33%) articles were graded as having high quality, 52 (30%) articles were graded as having considerable quality, 54 (31%) articles studies were graded as having moderate quality, and 5 (3%) articles were graded as having poor quality (Table 5). Five articles did not provide sufficient information to permit a complete MMAT appraisal and were graded as unclassified. See S3 Appendixfor detail of the quality assessment for each study.

**Table 5:** Distribution of MMAT scores (0 = lowest score and 100 = highest score)

| **MMAT Score distribution** | **Number of studies (%)** |
| --- | --- |
| 20 | 5 (3) |
| 40 | 17 (10) |
| 60 | 37 (21) |
| 80 | 52 (30) |
| 100 | 56 (33) |
| Total | 172 (100) |

## Discussion

This scoping review was conducted to map evidence on implementing the WHO’s essential lists in Africa to guide the effective implementation of the new WHO EDL. Our comprehensive scoping review identified themes based on the SURE framework into the barriers and enablers for implementing WHO essential lists across 172 articles. In lieu of the novelty of the EDL, there was limited published primary research on the implementation of WHO EDL. We found many studies reporting evidence on the implementation of EML in Africa. The review findings showed that the main theme barrier facing the more established EML, and newly introduced EDL was poorly equipped health facilities that entailed unavailability of essential in vitro diagnostics and medicines, stock-outs of laboratory reagents, and inadequate infrastructure and space to enable health service delivery. The EDL implementers at the national levels can work on equipping health facilities to improve the impact of the EDL.

Most of the studies in our review used quantitative methods, with nearly two-thirds of all studies using cross-sectional study designs in the surveys. There were fewer qualitative studies, mixed-methods studies, and randomized trials. Qualitative studies are useful for exploring and understanding barriers and facilitators for the EDL at context levels ^189^. Experimental trials are more useful for testing interventions that may improve the effectiveness of the EDL on health outcomes ^190^.

Similar work to ours, a systematic review by Peacocke et al., ^191^ explored the process of adapting the WHO EML at the national level. The authors provided key insights on the complexities and interdependencies essential to the implementation of the EML. Their review focused on key factors influencing the adaptation and implementation process of the EML at the macro level of the health system: country-level institutional structure; legislative and regulatory frameworks; governance, leadership and coordination for NEMLs. Our review provides further insights and maps evidence on implementing the WHO EDL and EML at national levels, focusing on the African context. In this review, we present barriers and enablers facing the EDL and EML at different levels of implementation; individual, health system, and social and political levels that influence the implementation of the WHO essential lists in-depth.

The essential lists and, more recently, the EDL by themselves are not sufficient to ensure their impact on access and health outcomes. A good health system is vital to strengthen the existence of the lists. Indeed, our review highlighted that health systems constraints remain the main barrier to implementing the EDL and EML. Such barriers included poorly equipped health facilities which entailed limited availability of essential tests and medicines, limited access to care facilities by patients due to financial and geographical constraints, availability of essential medicines and diagnostics at referral hospitals but not primary health care facilities, limited staff training, inefficient information systems, and inefficient procurement processes leading to regular stock- outs. In Africa, health systems face complex challenges such as the continued burden of communicable and non-communicable diseases pandemics amidst limited resources ^192–194^. In addition, many influencing factors in the health system determine the access, implementation, and effectiveness of diagnostic tests. Dealing with such challenges requires that decisions on health systems are informed by robust evidence that applies to the local context. Policymakers and health decision-makers can look to evidence-informed approaches, especially the synthesis of health policy and systems evidence, and contextualize findings to their settings. Methods for conducting or utilizing heath systems synthesis can be found in the WHO methods guide for evidence synthesis for health policy and systems ^195^.

Evidence-informed approaches are useful in guiding the adapting process and improving the implementation of the lists. WHO has a guidance resource for enabling African countries to adopt the WHO EDL to national contexts ^196^. To our knowledge, in Africa, Nigeria is the only country that has adapted the WHO EDL list and developed its own national EDL ^197^. Many African countries have adopted the WHO EML in national settings. However, stock-outs and limited access to medicines persist, emphasising the importance of enabling health systems to strengthen the implementation of the essential lists and ensure their impact. Evidence about the evidence-informed approaches or processes in adapting the EML has been published by South Africa ^93^, Ghana ^175^, and Tanzania ^66^. These publications highlighted enablers such as a well-structured and rigorous process ^93, 175^, utilization of evidence summaries in decision making ^93, 175^, involvement of a diverse committee and stakeholder engagements ^93, 175^. Challenges included insufficient and intermittent funding ^175^, limited use of scientific evidence ^66^, lack of expertise in evidence synthesis ^66, 175^ and health economic analyses ^66, 93^ in the review and development of NEML. Besides providing adaption guides for the essential lists, the implementation handbook guides can be released in conjunction with the versions of the model lists.

The WHO also released a handbook for monitoring the building blocks of health systems structured around the six main building blocks of WHO health systems framework’s: service delivery, health workforce, health information systems, access to essential medicines, financing, and leadership and governance ^198^. The proposed measures of health systems performance are crucial in health systems strengthening and valuable tools to accurately monitor the health system’s progress across the six building blocks over time. It facilitates the development of a sound Country monitoring strategy providing an enabling environment and sustainable scale-up of governance tools such as the EML and the newly introduced EDL. The EML and EDL play a vital role in realising UHC and access to quality health service delivery ^199^. The impact of the essential medicine and diagnostics lists will become truly effective only in well-functioning strengthened health systems. The core indicators to performance measures of key building blocks, including access to essential medicines and technologies, health service delivery, health workforce, health information systems, health financing, and leadership and governance ^198^, are all critical to the development, review, and implementation of the essential lists. The use of core indicators in the health systems could also assist in addressing EDL and EML implementation barriers timely, efficiently, and effectively to impact populations’ health outcomes.

In this review, there were notable successes of interventions developed to address barriers to the EML implementation that could be considered useful in the EDL implementation. The RDF ^92, 121, 200^, PBF ^119, 183, 184^, and P4P ^67, 182^ interventions addressed several barriers to implementing the EML: accessibility for care-related barriers, facility-related barriers, incentive-related barriers, information system-related barriers, accountability-related barriers, and facility financial resource-related barriers. The revolving fund pharmacy (RFP) ^201^, accredited drug dispensing outlets (ADDOS)^55^, and auditable pharmaceutical services and transaction system (APTS) ^84^ interventions also addressed the facility-related barriers. They contributed to the improved availability of essential medicines. Procurement and distribution-related barriers were addressed through direct distribution of supplies from partners ^50, 60, 130, 164, 167^, PBF ^62^, RDF programmes ^92, 121, 200^, and utilization of ICT ^68, 70^ in stock management. Similar interventions could be used to address the shortfalls of the EML and strengthen the EDL implementation designs. However, considering the country’s context and specificities to be addressed will be crucial when implementing interventions. Some interventions worked in some contexts and did not work in other contexts. For instance, the effect of PBF intervention did not affect the stock-out rate of essential medicines compared to payments not tied to the performance of essential medicines in some contexts ^164^. On the other hand, the provision of financial incentives in the P4P intervention addressed some of the health systems barriers; still, it was reported to have no evidence for increasing healthcare workers’ motivation ^182^. Though financial and non-financial incentives may motivate implementation, they can unrealistically raise expectations and hinder implementation in the long run due to sustainability issues ^202^.

We evaluated the existing literature through a systematic and rigorous process that involved reviewing qualitative, quantitative, and mixed methods studies using established guidance for scoping reviews. To inform the implementation of EDL, we also referred to a representative sample of the established EML. We did not include non-English studies hence could have missed studies published by French, Portuguese, or Arabic-speaking African countries. Secondly, due to accessibility limitations, we excluded 52 EML articles and multi-country studies about EML (n=67) due to the vast number of full-text EML studies and high likelihood of data saturation given rich, in-depth information from single countries in the multiple numbers of available studies. We also did not explore the process of adapting the WHO essential list to national contexts. Trend analysis from EML inception to the date of implementation aspects of the EML would help identify the successes, pitfalls, and plateaus of EML implementation over four decades. However, this was out of the scope of this work.

There has been limited primary research published on essential in vitro diagnostics in Africa since the introduction of the WHO EDL in 2018. Further studies can be conducted to provide contextual insights on the capacity of health systems to support the successful implementation of national EDLs bearing in mind the need to improve access to essential in vitro diagnostics in Africa. Consideration of dissemination and implementation frameworks such as the CFIR (Consolidated Framework for Implementation Research) ^203, 204^, RE-AIM (Reach, effectiveness, adoption, implementation, and maintenance) ^204, 205^, and PRISM (practical, robust implementation sustainability model) ^206, 207^ frameworks would be crucial when planning the implementation of the essential lists to guide adoption, adaptation, and evaluation of the lists. Qualitative research and process evaluations can be done to evaluate the impact of the essential lists and identify enablers and challenges to their implementation. More implementation trials or experimental studies can be conducted to assess effective interventions in different settings.

## Conclusion

The most dominant constraints facing EML implementation, a more established WHO essential list and the new EDL are mainly about the health system. The main theme barrier was poorly equipped health facilities, including limited availability of essential in vitro diagnostics and medicines and stock-outs, which mainly limit the implementation of the EML and EDL as well. The EDL implementation can learn from interventions to improve the availability and supply of essential medicines. When developing and implementing the National EDLs, consideration of these barriers will strengthen health service delivery, access to essential diagnostics and universal health coverage. Financial and non-financial incentives may be enablers, but their effect varies in different contexts.

## Supporting information

S1 Appendix_Deviations from the protocol

S1 Checklist_PRISMA checklist

S2 Appendix_Search strategy and SURE framework

S3 Appendix_Summary of included studies

## Data Availability

All data in this scoping review is included in the tables, figures and appendices.

## Acknowledgements

We thank Vittoria Lutje, Information Specialist at the Liverpool School of Tropical Medicine, who advised on and developed the search strategy. The authors express their gratitude to the Kenya Medical Research Institute for the support provided for this review.

## Contributors

MN, EO, and MM conceptualized and designed the study. MN, MM, BN, EJO, and EO contributed to the abstract and full article screening, data extraction, quality assessment, and synthesis of the included studies. MN prepared the first draft, and all authors critically reviewed the draft. EO is the senior researcher on the team, providing overall guidance.

## Funding

This study was funded by UK National Institute for Health Research (NIHR) Global HPSR developmental award (130222). NIHR has no role in the design, conduct, and interpretation of this review.

## Competing interests

None declared

## Supporting information

**S1 Appendix. Deviations from protocol.**

**S2 Appendix. Search strategy and SURE framework checklist.**

**S3 Appendix. Summary of included studies.**

**S1 Checklist. PRISMA-ScR checklist.**

**S1 Table. Barriers to the implementation of an essential diagnostic and medicines list.**

**S2 Table. Enablers for the implementation of an essential diagnostic and medicines list.**

## Notes

### Competing Interest Statement

The authors have declared no competing interest.

### Clinical Protocols

https://osf.io/cxwmu/

### Summary of Updates

We have revised the manuscript to remove line numbers to improve readability and remove errors in the document due to use of hyperlinks in some part of the manuscript.

