## Supplementary material for "Implementing essential diagnostics-learning from essential medicines: A scoping review": S1 Appendix_Deviations from the protocol

| **Section** | **Original protocol** | **Deviation from protocol method** | **Rationale** |
| --- | --- | --- | --- |
| Inclusion criteria | Inclusion of conference abstracts that have sufficient information on themes related to the implementation, uptake, and evaluation of the WHO essential lists (EDL&EML) | Exclusion of all conference abstract on essential medicines list (EML) | We excluded EML conference abstracts due to the vast number of full-text studies for the 40-year EML initiative and the high likelihood of data saturation. |
| Sampling | A convenience sample of  grey Literature will be accessed by searching for Global and National policy documents on EML and  EDL, diagnostic guidelines, and reports of ministry of health, health agencies through their websites  and links published in the last three years from selected African countries. | Omission of grey literature from data synthesis | For reasons of both scope and quality, our review restricted literature synthesis to included data from peer-reviewed journal articles from our electronic database search to ensure synthesis of most appropriate and rich data. |
