## Supplementary material for "Implementing essential diagnostics-learning from essential medicines: A scoping review": S2 Appendix_Search strategy and SURE framework

**MEDLINE Search Strategy**

Database: Ovid MEDLINE(R) and Epub Ahead of Print, In-Process, In-Data-Review & Other Non-Indexed Citations, Daily and Versions(R) <1946 to May 5, 2021>

Search Strategy:

--------------------------------------------------------------------------------

1	Drugs, Essential/ and World Health Organization/

2	WHO essential medicine list.mp.

3	WHO Model List of Essential Medicines.mp.

4	("WHO essential diagnostic list*" or WHO essential diagnostic or WHO Model List of Essential Medicines or WHO EML or WHO essential medicines).mp

5	World Health Organization essential medicine*.mp.

6	*Drugs, Essential/

7	(essential adj2 (drugs or medicines or diagnostic*)).mp.

8	1 or 2 or 3 or 4 or 5 or 6 or 7

**SURE framework for identifying considerations for the implementation of essential lists (EDL and EML)**

| **Level** | **Barriers and enablers** | **Description** |
| --- | --- | --- |
| Recipients of care | Knowledge and skills | Recipients of care may have varying degrees of knowledge about the healthcare issue or the intervention, or may not have the skills to apply this knowledge. E.g. People may be unaware that family planning services are available at their local clinic or may not have the skills to prepare oral rehydration therapy when its use has been recommended. |
|  | Attitudes regarding programme acceptability, appropriateness and credibility | Recipients of care may have opinions about the healthcare issue and the intervention, including views about the acceptability and appropriateness of the intervention and the credibility of the provider and the healthcare system. E.g. People may not agree with the choice of intervention or may not trust the reasons behind it |
|  | Motivation to change or adopt new behaviour | Recipients of care may have varying degrees of motivation to change behaviour or adopt new behaviours. E.g. they may be more or less motivated to seek care |
| Providers of care | Knowledge and skills | Providers may have varying degrees of knowledge about the healthcare issue or the intervention, or may not have the skills to apply this knowledge. E.g. health workers may be unaware of guidelines on tuberculosis treatment or may not have received training in the implementation of these guidelines |
|  | Attitudes regarding programme acceptability, appropriateness and credibility | Providers may have opinions about the healthcare issue and the intervention, including views about the acceptability and appropriateness of the intervention and the credibility of the provider and the healthcare system. E.g. health workers may not agree with the choice of intervention or may not trust the reasons behind it |
|  | Motivation to change or adopt new behaviour | Providers may have varying degrees of motivation to change behaviour or adopt new behaviours. E.g., they may be more or less motivated to take on new tasks |
| Other stakeholders (including other healthcare providers, community health committees, community leaders, programme managers, donors, policy makers and opinion leaders) | Knowledge and skills | Other stakeholders may have varying degrees of knowledge about the healthcare issue or the intervention, or may not have the skills to apply this knowledge. E.g. a community leader may have insufficient knowledge of the benefits of exclusive breastfeeding or may not feel skilled in running community meetings to promote infant care |
|  | Attitudes regarding programme acceptability, appropriateness and credibility | Other stakeholders’ may have opinions about the healthcare issue or the intervention, including views about the acceptability and appropriateness of the intervention and the credibility of the provider and the healthcare system. E.g. stakeholders may not agree with the choice of intervention because of competing interests or priorities |
|  | Motivation to change or adopt new behaviour | Other stakeholders may have varying degrees of motivation to change behaviour or adopt new behaviours. E.g. programme managers may not be motivated to deliver supervision to remote clinics |
| Health system constraints | Accessibility of care | The accessibility of healthcare facilities may affect implementation of the option, for instance because of financial (user fees), geographic (distance to clinic), or social (access for certain ethnic groups) factors |
|  | Financial resources | Additional financial resources may be needed to implement the option |
|  | Human resources | An increased supply or distribution of health workers may be needed to implement the option |
|  | Educational system | The educational system for health workers may need to be modified |
|  | Clinical supervision | Health workers may require more supervision than is currently provided to implement the option |
|  | Internal communication | Changes in communication between different levels of the health system or between the health and social care systems may be needed to implement the option |
|  | External communication | Changes in communication between health workers and recipients of care needs may be needed to implement the option |
|  | Allocation of authority | Changes may be needed regarding the levels or individuals that have the authority to make decisions |
|  | Accountability | Changes may be needed so that those with the authority to make decisions are accountable for the decisions they make |
|  | Management and or leadership | Adequately trained managers or sufficient leadership may be needed to implement the option |
|  | Information systems | Adequate information systems to assess and monitor needs, resource use, and utilisation of targeted services may be needed to implement the option |
|  | Facilities | Adequate supply and distribution of necessary supplies and equipment to facilities, and maintenance of these facilities, may be needed to implement the option |
|  | Patient flow processes | Adequate processes for outreach and receiving, referring and transferring patients may be needed to implement the option |
|  | Procurement and distribution systems | Adequate systems for procuring and distributing drugs and other supplies may be needed to implement the option |
|  | Incentives | Reimbursement systems for patients, health workers or others may need to be structured to facilitate rather than hinder implementation of the option |
|  | Bureaucracy | Paperwork and procedures may need to be structured to facilitate rather than hinder implementation of the option |
|  | Relationship with norms and standards | Current norms and standards of practice need to be in line with the relevant option |
| Social and political constraints | Ideology | Ideological beliefs (e.g. in ‘free markets’) may affect implementation of the option |
|  | Short-term thinking | Implementation of the option may be opposed if its benefits are likely to occur beyond the time horizon of decision makers (e.g. after the next election) |
|  | Contracts | Contracts with service providers or enforcement of contracts may not be adequate to ensure implementation of the option or the types of effective care at which it is targeted |
|  | Legislation or regulations | Changes to legislation or regulations, including those that are general (e.g. regulating government contracts, regulating working conditions) and those that are specific to the health system (e.g. licensing health professionals) may be needed |
|  | Donor policies | Donor policies and programmes may influence implementation |
|  | Influential people | The opinions of influential people may influence the option or the types of effective care at which it is targeted |
|  | Corruption | Corrupt behaviour by decision makers or others may influence implementation |
|  | Political stability | Political instability may influence implementation |
