## Supplementary material for "Implementing essential diagnostics-learning from essential medicines: A scoping review": S3 Appendix_Summary of included studies

|  | **Author and year of publication** | **Topic Area** | **Aims** | **Study design** | **Participants** | **Context** | **Type of facility** | **Key findings** | **MMAT SCORE** |
| --- | --- | --- | --- | --- | --- | --- | --- | --- | --- |
| 1. | Shumbej T et al;2020 | EDL | Assess essential in-vitro laboratory service provision in accordance with the WHO standards in Guragae Zone primary health care unit level, South Ethiopia. | Cross-sectional study | Other | Ethiopia | Primary Health care facilities | Generally, the findings indicate that major gaps regarding EDL, limited items of the major laboratory equipment, and stock problems on maintaining main supplies for essential laboratory services provision. | 100 |
| 2. | Ward C et al; 2021 | EDL | To determine the availability and pricing of laboratory testing in the Northern Region of Ghana to identify current gaps with respect to the WHO's Essential Diagnostics List | Cross-sectional study | Health care workers | Ghana | Primary Health care facilities; Laboratories | Data on accessibility of essential in vitro diagnostics in community-based health planning and services (CHPS) sites. | 100 |
| 3. | Wahlfeld C. et al; 2019 | EDL | To evaluate the inventory levels of HIV RDT kits at specific healthcare facilities in Zambezia province, Mozambique | Cross-sectional study | Other | Mozambique | Primary Health care facilities | Differences in inventory levels among the districts in the Zambezi province. | 60 |
| 4. | Horton S et al; 2019 | EDL | To compare the most common diagnostic/laboratory tests across five different referral hospitals by  volume and revenue | Cross-sectional study | Other | Kenya, Nigeria | Hospitals | The challenge remains to ensure that these EDL tests will be affordable in relation to price of the tests in different facilities. | 80 |
| 5. | Shiferaw F. et al; 2018 | EDL & EML | To highlight the burden of NCDs in Ethiopia and analyzing one of the two major WHO recommended policy issues | Mixed methods study | Other | Ethiopia | Unclear | Data availability on NCD morbidity and mortality is limited. Shortage of essential NCD drugs and diagnostics facilities and lack of treatment guidelines. | 80 |
| 6. | Isadru V et al; 2021 | EDL & EML | This study sought to determine the readiness of HFs in managing hypertension (HTN) and diabetes cases at primary health facilities in the Bidibidi refugee settlement, Yumbe district, Uganda. | Cross-sectional study | Health care workers | Uganda | Primary Health care facilities | Availability of diagnostics conducted on-site was 96%, availability of essential drugs was low at 58.8%, 18.8% of the HFs had all the 5 essential drugs assessed, and 43.8% had 1-2 of the assessed essential drugs available at the time of assessment. | 100 |
| 7. | Ndagire E et al; 2021 | EDL & EML | To assess health system gaps in delivery of rheumatic heart disease related care in Uganda | Mixed methods study | Health care workers | Uganda | Hospitals; Primary Health care facilities | Unavailability of diagnostics tests for rheumatic fever and rheumatic heart disease in many districts | 80 |
| 8. | Bay N et al; 2019 | EDL & EML | Assess the management of hypertension in an emergency department (ED). | Cross-sectional study | Patients; Health care workers | Mozambique | Hospitals | Stock-outs of basic diagnostic tools for risk stratification and medicines were registered. The availability of medicines was 28% on average. | 100 |
| 9. | Elnour F et al; 2019 | EDL & EML | To evaluate the management of severe malaria at Gezira State hospital in Sudan by assessing hospital readiness, health care provider knowledge and the care received by severe malaria patients | Cross-sectional study | Patients; Health care workers | Sudan | Hospitals | This study revealed that management of severe malaria at the hospital level was substandard.  From all hospitals evaluated, 90% (18/20) were suffering from shortage of staff, especially doctors. About half of the health care providers (46.7%) did not receive severe malaria management training. Microscopy was available in all hospitals (100%), while rapid diagnostic test, complete blood count and renal function test were available in 15 hospitals (75%). | 80 |
| 10. | Jessen N et al; 2018 | EDL & EML | To evaluate access (availability, prices, and affordability) to CVD medicines and diagnostics in the public and private healthcare sectors, in Maputo City, Mozambique. | Cross-sectional study | Health care workers | Mozambique | Hospitals; Pharmacies/Dispensaries | Availability of essential CVD medicines is below the 80% target in both the public and private sectors. | 40 |
| 11. | Webb E.M. et al; 2019 | EDL & EML | To assess the capacity of primary care clinics in one district to  provide quality diabetes care. | Cross-sectional study | Health care workers | South Africa | Hospitals | The capacity to deliver quality care is compromised by the poor availability of guidelines, educational material, and the absence of monofilaments. | 40 |
| 12. | Gonzo. et al; 2020 | EDL & EML | To assess the adequacy of IDA diagnosis and management within the Namibian private healthcare system. | Cross-sectional study | Health care workers | Namibia | Hospitals; Laboratories | Medical practitioners follow different guidelines and reference documents when dealing with patients with suspected. | 100 |
| 13. | Rogers H et al; 2018 | EDL & EML | To conduct a needs assessment in higher-level public sector facilities capacity to detect and manage NCDs, | Cross-sectional study | Health care workers | Uganda | Hospitals; Primary Health care facilities | All facilities reported deficiencies in NCD screening and management services. | 40 |
| 14. | Tickell K.D. et al; 2019 | EDL & EML | To determine adherence to pediatric inpatients care recommendations | Mixed methods | Health care workers; Laboratory workers | Kenya, Uganda, Malawi, Burkina Faso | Hospitals | All clinical laboratories have the infrastructure to provide basic services, however, reagent stock outs intermittently disrupted services in certain sites. Essential medicines were largely available during assessment visits, but frequent stockouts were reported. | 60 |
| 15. | Rawlins B et al; 2018 | EDL & EML | To analyze and present findings on the quality of clinical practices for the identification and management of PE/E during antenatal care (ANC) and labor and delivery. | Cross-sectional study | Patients; Health care workers; Health care managers | Ethiopia, Kenya, Madagascar, Mozambique, Rwanda, and Tanzania, including Zanzibar, | Hospitals;Primary Health care facilities;Pharmacies/Dispensaries | The observed ANC consultations showed a low level of provision of WHO-recommended PE/E screening practices in ANC services with only one third (31%) of the ANC clients receiving the recommended screening | 100 |
| 16. | Martei Y et al; 2019 | EML | To measure the impact of cancer medicine stock outs on delivery of optimal therapy in Botswana. | Cohort study | Patients | Botswana | Hospitals | Our study identified a high rate of cancer medicine stock outs affecting standard regimens for commonly treated cancers in Botswana | 60 |
| 17. | Kuwawenaruwa A et al; 2019 | EML | To assess the effects of medicine availability and stock-outs on healthcare utilization in Dodoma region, Tanzania. | Cross-sectional study | Community residents; Health care workers | Tanzania | Hospitals; Primary Health care facilities; Pharmacies/Dispensaries | The availability of most tracer medicines was relatively good, with continuous availability | 60 |
| 18. | Tsofa B. et al; 2017 | EML | To analyse the early implementation experiences of human Resources for Health (HRH) and Essential Medicines and Medical Supplies (EMMS) management. | Qualitative study | Health care workers; Health care managers | Kenya | Local health managers | The human resources for health and essential medicines and medical supplies management functions were rapidly transferred to counties before appropriate county-level structures and adequate capacity were in place. | 100 |
| 19. | Brault M et al; 2017 | EML | To evaluate national policy and strategy documents, quantitative indicator data, and qualitative data to identify barriers and facilitators influencing child survival in Kenya. | Qualitative study | Patients; Health care workers; Laboratory workers; Health care managers; National Essential List Committees; Other | Kenya | Hospitals; Community residents | Free care was a major strength of the healthcare system that improved access to and utilization of MNCH services by enabling low-income families to obtain care more easily. Stockouts of essential medicines and commodities were common across facilities. | 100 |
| 20. | Sieleunou I. et al; 2019 | EML | To examine the effects of Performance based Financing on the availability of essential medicines (EMs) in low- and middle-income countries. | Qualitative study | Health care workers | Cameroon | Primary Health care facilities | Performance Based Financing intervention improved the perceived availability of EMs in three regions in Cameroon. | 80 |
| 21. | Perumal-Pillay V et al; 2016 | EML | To compare latest SA STG/EMLs with the latest World Health Organization (WHO) Model EMLs to assess or evaluate alignment of these lists. | Observational, descriptive study design | Other | South Africa | Hospitals; Primary Health care facilities | There have been many additions and deletions of medicines to the STG/EMLs over time. Possible reasons for deletions include safety and efficacy reasons; adverse effects and/or the availability of safer or cheaper alternatives. | 80 |
| 22. | Kibuule D et al; 2016 | EML | To evaluate public sector pharmaceutical policies and guidelines influencing the therapeutic use of Cotrimoxazole, Amoxicillin and Azithromycin (CAA) antibiotics in Namibia. | Quantitative policy analysis | Other | Namibia | Ministry of Health and Social Services | Despite the existence of guidelines for antibiotic use in various treatment guidelines in Namibia including HIV/AIDS, malaria and tuberculosis as well as the integrated management of childhood illnesses in Namibia, Namibia has no comprehensive antibiotic policy, antibiotic guidelines, antibiotic formulary or antibiograms. | N/A |
| 23. | Khan R. et al; 2019 | EML | To strengthen provision of essential medicine to women and children in post-Ebola in Sierra Leone. | Interrupted Time Series Analysis | Other | Sierra Leone | Hospitals | This program resulted in 100% availability of essential medicines at KGH during the ten-month implementation phase, which has been maintained to date. The increased human resources, decentralized drug distribution systems and robust data collection systems enabled the MOHS to provide approximately 80% of medications, with PIH filling the gap for the remaining 20%. | N/A |
| 24. | Anselmi L et al; 2017 | EML | To test the causal pathways through which P4P schemes may (or may not) influence maternal care outcomes. | Interrupted Time Series Analysis | Patients; Health care workers | Tanzania | Hospitals; Primary Health care facilities; Pharmacies/Dispensaries | P4P led to an increased availability of resources at the facility, notably a reduction in the disruption of services due to broken equipment, a reduction in the stock-out rate of essential medical and drugs particularly those used during delivery. | 80 |
| 25. | Lillian RR. et al; 2018 | EML | To perform an evaluation of primary eye care services in three districts of South Africa. | Cross-sectional study | Health care workers | South Africa | Hospitals | The study demonstrated poor levels of primary eye care services, with multiple challenges in organisation of care, availability of essential resources and clinical knowledge of primary health care workers. | 40 |
| 26. | Tumwine Y et al; 2010 | EML | To assess the impact of the Pull system on the availability and reduction of expiry of essential medicines and medical supplies and to determine factors affecting their availability in Kilembe Hospital, Uganda. | Cross-sectional study | Health care workers;Health care managers;Other | Uganda | Hospitals | The median number of days out of stock for drugs and medical supplies was higher in the push system compared to the pull system. Inadequate training, lack of transport and inadequate funding affecting availability of essential medicines. | 40 |
| 27. | Richard O.T. et al; 2015 | EML | To examine the major challenges affecting the hybrid system of medicines supply. | Cross-sectional study | Health care workers | Uganda | Primary Health care facilities | Collaborative linkages with the National Medicines Stores (NMS) is weak and quantification of essential medicines by health workers under the hybrid system is poor. | 60 |
| 28. | Masembe I.K. et al; 2016 | EML | The main purpose of this study was to analyse the effect of strategic planning on procurement of medicals in Uganda’s regional referral hospitals | Mixed methods study | Other | Uganda | Hospitals | Strategic planning was affected by funding. The quantity and some extent of the quality of medicals to be procured was determined by the amount of funds available to the RRH. | 20 |
| 29. | Mikkelsen-Lopez.I. et al 2013 | EML | Assess whether reform in the Tanzanian medicines delivery system from a central push kit system to a decentralized pull Integrated Logistics System (ILS) has improved medicines accountability. | Case study | Health care workers | Tanzania | Hospitals;Primary Health care facilities | The Integrated Logistics System has not adequately addressed accountability concerns seen under the kit system due to a combination of governance and system-design challenges. | 80 |
| 30. | Rockers P et al; 2019 | EML | To evaluate the effect of Novartis Access on availability and price of non communicable disease medicines in eight counties in Kenya | Cluster-randomized controlled trial | Patients; Health care workers | Kenya | Hospitals;Primary Health care facilities | After 15 months, the programme had a positive effect on availability of amlodipine and metformin at facilities, but had no effect on availability of the other medicines in the Novartis Access portfolio. | 80 |
| 31. | Rutta E et al; 2015 | EML | The program’s goal was to improve access to affordable, quality medicines and pharmaceutical services in retail drug outlets in rural or peri-urban areas with few or no registered pharmacies. | Case study | Patients;Health care workers | Tanzania | Pharmacies/Dispensaries | The critical element that was essential to the ADDO program’s success is stakeholder engagement, the successful buy-in and sustained commitment came directly from the effort, time and resources spent to fully connect with vital stakeholders at all levels. | 40 |
| 32. | Sanogo NA et al; 2020 | EML | To explore the impact of a social health insurance scheme on the quality of antenatal care in Gabon | Qualitative study | Patients;Health care workers | Gabon | Hospitals;Primary Health care facilities | The mandatory health insurance of the NFHISG has improved accessibility of care and improving certain components of care provision. | 100 |
| 33. | Vledder M et al; 2019 | EML | To identify a nationally scalable structure for medicines distribution which leads to systematic improvement in medicines availability at the health facility level. | Randomized controlled trial | Health care workers | Zambia | Hospitals;Primary Health care facilities | Availability of essential medicines improved remarkably at the health facility level, especially wherein the MODEL B districts (directly to MSL). Higher gains in reduction of stockout rates were observed in MODEL B districts compared to the MODEL A districts (district stores). | 80 |
| 34. | Nass S.S et al; 2019 | EML | To measure the impact of integrative supportive supervision on the quality of primary health-care delivery. (on selected Katsina state, Nigeria) | Cross-sectional study | Health care workers | Nigeria | Primary Health care facilities | A steady increase in the availability of essential drugs among the health facilities from an average of 70-83%. an increase in availability from visit 2 among 13 health facilities found with less than 70% availability of essential drugs at the first visit. | 40 |
| 35. | Babigumira JB et al; 2017 | EML | To evaluate the impact of the training and deployment of pharmacy assistants to rural health centers in Malawi. | Quasi-Experimental design | Patients;Health care workers | Malawi | Primary Health care facilities | Pharmacy assistant training and deployment to rural health centers in Malawi increased access to antimalarial medications for children under five years in the short term but this effect was attenuated over the subsequent year. | 80 |
| 36. | Sakyi E et al; 2016 | EML | To investigate the opinions of health managers about the problems emanating from the national health insurance policy for hospital managers in regard to reimbursement, claims management, service delivery and waiting time. | Qualitative study | Health care managers | Ghana | Hospitals | The findings demonstrate that a major obstacle to effective healthcare delivery is irregular and unpredictable reimbursement patterns. On average, hospitals are reimbursed within ten weeks. | 100 |
| 37. | Manji I et al; 2017 | EML | To describe the utilization of the RFP model within the AMPATH catchment area | Interrupted time series | Health care workers | Kenya | Primary Health care facilities | RFP model substantially and sustainably improved access to healthcare to patients in both rural and urban selfing and access to essential medicines through a backup of the MOH pharmacies. | 60 |
| 38. | Beyene D et al; 2020 | EML | To assess the outcome performance of pharmaceuticals services among selected hospitals with and without the Auditable Pharmaceutical and Transaction System (APTS) systems in SNNPR, Ethiopia | Cross-sectional comparative facility-based study | Patients;Health care workers;Health care managers | Ethiopia | Hospitals | APTS implementation has a positive effect; on patient knowledge of correct dosage and satisfaction, and positive impact the mean availability of Essential drugs at stores. | 60 |
| 39. | Binyaruka P et al; 2017 | EML | To evaluate the effects of payment for performance (P4P) on the availability and stockout rate of reproductive, maternal, newborn and child health (RMNCH) medical commodities in Tanzania and assess the distributional effects. | Interrupted Time Series Analysis | Health care workers;Health care managers;Community residents | Tanzania | Hospitals;Primary Health care facilities;Pharmacies/Dispensaries | P4P was associated with an 8.4 percentage point increase in the availability of all 37 medicines combined and an 8.3 percentage point increase in the availability of medical supplies. | 40 |
| 40. | Sieleuonou I et al; 2018 | EML | To examine the effects of the PBF intervention on the availability of essential medicines (EM). | Randomized controlled trial | Health care workers;Health care managers | Cameroon | Primary Health care facilities | The PBF intervention had no effect on stock-outs of antenatal care drugs, vaccines, IMCI drugs, and labour and delivery drugs. However, the intervention was associated with a significant reduction in stock outs of family planning drugs. | 20 |
| 41. | Agodokpessi G et al; 2015 | EML | The evaluation of the operation of the revolving drug fund (RDF) | Cross-sectional study | Health care workers | Benin | Hospitals | The RDF provided ICS at affordable prices for the majority of asthma patients. For a small minority of poor asthma patients who could afford to pay, the ICS were provided at no cost. | 40 |

| 42. | Kibirige D et al;2017 | EML | To provide contemporary information about the availability, cost, and affordability of medicines and diagnostic tests integral in the management of DM and CVD in Uganda | Cross-sectional study | Health care workers | Uganda | Hospitals;Pharmacies/Dispensaries | The majority of medicines and diagnostic tests important in DM and CVD care are largely unavailable and unaffordable in Uganda. | 100 |
| --- | --- | --- | --- | --- | --- | --- | --- | --- | --- |
| 43. | Bintabara D et al 2019 | EML | Assess the availability and readiness of health facilities to provide basic emergency obstetric and newborn care (BEmONC) and its associated factors. | Cross-sectional study | Health care workers | Tanzania | Hospitals;Primary Health care facilities;Pharmacies/Dispensaries | There are disparities in the availability and readiness to provide BEmONC among health facilities in Tanzania. Overall, availability of seven signal functions and average readiness score were consistently higher among hospitals than health centers and dispensaries. | 100 |
| 44. | Lonnée HA  et al 2021 | EML | Assess the type of anaesthesia used for caesarean sections in Sierra Leone. | Cross-sectional study | Health care workers | Sierra Leone | Hospitals | The supply of analgesics was poor, with oral paracetamol, nonsteroidal anti-inflammatory drugs (NSAIDs) and tramadol always available at 90%, 90% and 94% of hospitals, respectively. | 100 |
| 45. | Orji I.A. et al 2021 | EML | Assessment of capacity and readiness for a system-level hypertension control program within the Federal Capital Territory of Nigeria. | Cross-sectional study | Health care workers | Nigeria | Primary Health care facilities | Demonstrated the feasibility of implementing the hypertension treatment in Nigeria program based on the workforce, equipment and paper-based information systems, but identified critical needs for health worker training, protocol implementation, and essential medicine supply strengthening for hypertension treatment and control. | 100 |
| 46. | Surgey G et al 2019 | EML | Describe the journey of the introduction of HTA into decision-making processes through a case study revising the National Essential Medicines List (NEMLIT) in Tanzania | Situational analysis | National Essential List Committees | Tanzania | Unclear/Not reported | In Tanzania, it has been recognized that HTA can play a critical role in an efficient healthcare system. Cost-effectiveness has been considered in the development of the EML and adherence to the STG/NEMLIT is being enforced by the Ministry. | N/A |
| 47. | Perumal-Pillay V.A et al;2020 | EML | Improve the understanding of how formulary managers of selected medical schemes made decisions for the selection of medicines for their formularies. | Qualitative study | Health care stakeholders | South Africa | Health care stakeholders | The process of and criteria for medicines selection and evidence based review of formularies were also similar. Selection of medicines was inherent in the formulary review process | 80 |
| 48. | Muthathi I.S. et al; 2020 | EML | To analyse PHC facility managers’s decision space and their participation in the implementation of the Ideal Clinic Realisation and Maintenance (ICRM) programme. | Cross-sectional study | Health care managers | South Africa | Primary Health care facilities | PHC facility managers reported lack of involvement in the conceptualization of the ICRM programme | 80 |
| 49. | Hunie M et al;2020 | EML | Assess the availability of emergency drugs and essential equipment in ICUs in hospitals in Ethiopia. | Cross-sectional study | Health care workers | Ethiopia | Hospitals | The availability of essential emergency equipment and drugs was found to be generally inadequate. | 60 |
| 50. | Dzudie A et al;2020 | EML | Assess the availability, cost, and affordability of essential cardiovascular medicines in the South-West region of Cameroon. | Cross-sectional study | Health care workers | Cameroon | Pharmacies/Dispensaries | Overall, the mean availability of essential medicines for CVD was 33%, much lower than the 80% recommended by the WHO Global Action Plan for Prevention of NCD. | 100 |
| 51. | Tadesse T et al; 2021 | EML | Assessing the availability and affordability of essential medicine for children in selected health facilities of southern nations, nationalities, and peoples‚ regions (SNNPR), Ethiopia. | Cross-sectional study | Health care workers | Ethiopia | Hospitals;Primary Health care facilities;Pharmacies/Dispensaries | The availability varied by sector, type of medication, and level of health facilities. The average availability of essential medicines was 57% for the public sector and 53% for the private sector. | 80 |
| 52. | Mukundiyukuri J.P. et al;2020 | EML | This study described the availability, costs, and stock-outs of essential NCD drugs in three rural Rwandan districts | Cross-sectional study | Health care workers | Rwanda | Hospitals | The high availability of NCD drugs in the study is attributed to integrations of NCD care into primary health care. | 60 |
| 53. | Kirua R.B. et al; 2020 | EML | To compare the prices of medicines used in the management of pain, diabetes, and cardiovascular diseases in private pharmacies and the National Health Insurance Fund (NHIF) in Tanzania. | Cross-sectional study | Health care workers | Tanzania | Pharmacies/Dispensaries | There is a large variation in prices of surveyed essential medicines sold in private pharmacies in the sampled four regions in Tanzania. | 80 |
| 54. | Armstrong-Hough M et al; 2020 | EML | Analysis of the availability and cost of a selection of essential medicines for NCDs in a sample of Ugandan health facilities over a five-week period | Cross-sectional study | Health care workers | Uganda | Hospitals;Primary Health care facilities;Community services | Availability of medicines varied substantially over time, especially among public facilities. Among private-for-profit facilities, the cost of the same medicine varied from week to week. Private-not-for-profit facilities experienced less dramatic fluctuations in price. | 60 |
| 55. | Bravo M.P. et al; 2019 | EML | Support of HIV and tuberculosis (TB) testing and treatment supported by PEPFAR in Africa requires immense quantities of tests and medications. | Longitudinal analysis | Health care workers | Mozambique | Primary Health care facilities;Pharmacies/Dispensaries | There were improvements in medication availability over time, even in the face of burgeoning demands from new policies. Periodic medication stock outs were also documented. | 40 |
| 56. | Wood-Thompson D.K et al; 2018 | EML | To determine current sedation practices, common indications, and major obstacles in selected emergency centres across Southern Gauteng, South Africa, with a view to improving future standards and practices. | Cross-sectional study | Health care workers;Health care managers | South Africa | Hospitals | The majority of centres performed between 10 and 30 procedures per month requiring sedation. Staff training in the practice of procedural sedation was mostly obtained internall. Essential drugs, procedure monitors, resuscitation equipment and protocols were all available in 70% of centres. | 60 |
| 57. | Ayenew A. et al; 2020 | EML | To determine essential newborn care associated factors among obstetrical care providers in Awi zone Health facilities. | Cross-sectional study | Health care workers | Ethiopia | Primary Health care facilities | Having in-service training, midwifery profession, a good knowledge of essential newborn care, availability of drugs, level of education and availability of medical equipment for essential newborn care were the determinant factors for essential newborn care practice. | 80 |
| 58. | Gebremedhin T. et al; 2019 | EML | To evaluate the process of the CBNC program implementation in Geze Gofa district, south Ethiopia | Mixed methods study | Patients;Health care workers;Health care managers | Ethiopia | Primary Health care facilities | The overall process of the community-based newborn care program implementation was 72.7%, as measured by the availability of medical equipment and drugs, healthcare providers compliance with the national guideline and maternal satisfaction. | 100 |
| 59. | Wirtz V.J et al; 2017 | EML | To assess access to Noncommunicable diseases medicines in Kenya for patients diagnosed and prescribed treatment for asthma, diabetes and hypertension. | Cluster randomized controlled evaluation | Patients;Health care workers | Kenya | Health sector | Majority of patients purchased medicine in private sector in particular patients with asthma because asthma medication are likely to be available in public facilities. | 80 |
| 60. | Sumankuro J. et al; 2018 | EML | To examine the health system factors impacting on access to and delivery of quality maternal and newborn healthcare in rural settings. | Qualitative study | Community residents;Health care workers | Ghana | Hospitals;Primary Health care facilities | Inadequate medical equipment and essential medicines, infrastructural challenges, shortage of skilled staff, high informal costs of essential medicines and general limited capacities to provide care were significant barriers affecting the quality and appropriateness of maternal and neonatal healthcare. | 100 |
| 61. | Andriantsimietry S et al; 2016 | EML | To assess service availability and readiness assessment of Maternal, Newborn and Child Health Services at Public Health Facilities in Madagascar. | Cross-sectional study | Health care workers | Madagascar | Hospitals;Primary Health care facilities | Availability of essential diagnostics in antenatal clinics varied depending on the level of hospital with Urine dipstick protein was 100% available in university hospitals, 85.7% in referral district and regional hospital, and 5% in basic health centres; Hemoglobin test was 83.3% available in university hospitals | 60 |
| 62. | Mugo N et al; 2018 | EML | To understand the community members‚experience, perceptions, and the barriers in relation to accessing and utilizing maternal healthcare services in South Sudan | Qualitative study | Patients | Sudan | Hospitals;Primary Health care facilities | Inadequate quality of antenatal care services such as lack of essential medicine, supplies and tools was linked to individuals’ mothers’ dissatisfaction with the services received. | 60 |
| 63. | Mutale W. et al; 2018 | EML | To assess Zambian health system s capacity to address NCD using an adapted WHO Essential Non Communicable Disease Interventions tool. | Cross-sectional study | Health care workers | Zambia | Hospitals;Primary Health care facilities | Nearly all primary health facilities studies could not meet minimum threshold to manage NCD in line with WHO recommendations. | 100 |
| 64. | Millogo O et al; 2021 | EML | To determine the association between health facility malaria services readiness and Malaria mortality in children under the age of 5 years in Burkina Faso | Cross-sectional study | Health care workers | Burkina Faso | Hospitals;Primary Health care facilities;Pharmacies/Dispensaries | Malaria mortality rate in medical centres was 4.8 times higher than that of peripheral health centres. Essential medicines were the domain with the lowest readiness. Higher readiness index was associated with lower mortality in peripheral health centres. | 80 |
| 65. | Ngcobo N.J. et al; 2017 | EML | To establish the status of vaccine availability and associated factors in government health facilities of Tshwane Health District in Gauteng Province, SA | Cross-sectional study | Health care workers;Health care managers;Other | South Africa | Primary Health care facilities | Vaccine shortage occur in Tshwane clinics and last for up to more than 2 weeks. | 60 |
| 66. | Orubu, E. et al; 2016 | EML | Examined compounding records from January to December 2011 in a sample of seven hospitals to describe what medicines for oral use were commonly compounded in Nigeria | Retrospective descriptive study | Health care workers | Nigeria | Hospitals | The main reason for compounding was the unavailability of age-appropriate formulations. Comprehensive stability testing was not reported for the products. | 80 |
| 67. | Oridanigo E. et al; 2021 | EML | To assess the perceived affordability of essential medicines and associated factors in public health facilities of the Jimma Zone, Southwest Ethiopia. | Cross-sectional study | Health care workers | Ethiopia | Hospitals;Primary Health care facilities | Selected key essential medicines used for the treatment of common health problems were not affordable in the surveyed health facilities. | 80 |
| 68. | Magadzire B.P. et al; 2017 | EML | To identify the causes of stock-outs and to illustrate how they undermine access to medicines (ATM) in the Western Cape Province, SA. | Qualitative study | Health care workers;Health care managers | South Africa | Primary Health care facilities | Stock-outs resulted from the following inefficiencies at the central role. Stock-outs undermined the intended benefits of ATM strategies. | 100 |
| 69. | Niankara I. et al; 2018 | EML | To model the joint impact of generic essential drugs and nursing staff supplies constraints on access to primary health care in the country. | Cross-sectional study | Other | Burkina Faso | Primary Health care facilities | Shortages in nursing staff supply do put a significant constraint on access to primary health care. | 40 |
| 70. | Bekele A. et al; 2017 | EML | To assess service availability and readiness for diabetes health care | Cross-sectional study | Health care workers | Ethiopia | Hospitals;Primary Health care facilities;Other | Among all health facilities that offer services for non-communicable diseases, 59 % offer services for diabetes. Availability of medicines for diabetes varies by facility type, and is higher in hospitals when compared with other facility types (ranging from 90 % in hospitals to 0 % in clinics). | 100 |
| 71. | Kabunga L et al; 2017 | EML | To understand the relationship, between access to credit services through financial loans or stock and availability of essential child medicines and licensing status among medicine retail outlet including drug shops and pharmacies. | Cross-sectional study | Health care workers | Uganda | Pharmacies/Dispensaries | For all essential child medicines assessed, access to credit through financial loans through obtaining stock on credit did not influence stock availability. Access to credit services through loans or through stock on credit was seen to influence licencing status. | 100 |
| 72. | Mukose A.D. et al; 2019 | EML | To explore health providers experiences of preparedness and organization of Option B + services in Central Uganda. | Qualitative study | Health care workers | Uganda | Primary Health care facilities | Health providers and facilities were reasonably prepared before and during the implementation of option B+. Health facilities were stocked with necessary essential medicines and health supplies such as ARV medications, HIV and EID testing kits. | 100 |
| 73. | Kuwawenaruwa A. et al; 2020 | EML | Analyses accountability mechanisms contributing to the performance of Jazia PVS in Tanzania. | Qualitative study | Health care workers;Health care managers;Other | Tanzania | Hospitals;Primary Health care facilities | Financial, performance, and procedure accountability measures played an important role for the successful performance of Jazia PVS in Tanzania | 100 |
| 74. | Saiyoki L. et al; 2019 | EML | To determine accessibility to essential medicines for the four major non communicable in Trans-Nzoia County. | Cross-sectional study | Health care workers | Kenya | Hospitals | Majority of the participants at  71% felt that the county government was not doing enough to ensure steady supply of the essential medicines for noncommunicable diseases. | 60 |
| 75. | Legesse H. et al; 2019 | EML | To present challenges, strategies in availing lifesaving commodities at health posts and recommendation. | Cross-sectional study | Health care workers | Ethiopia | Primary Health care facilities | The current supply chain is not strong to sustainably avail lifesaving commodities at health-posts | 40 |
| 76. | Koduah A. et al; 2019 | EML | To examine the 2017 STGs and EML review process, the actors involved and how the list of medicines and disease conditions evolved between the last two editions. | Case study | Other | Ghana | Healthcare stakeholders | Ghana’s experience in using evidence from research and practice and engaging wide stakeholders can serve as lessons for other low and middle-income countries. | N/A |
| 77. | Brenner S. et al; 2017 | EML | To evaluate the impact of a performance-based financing scheme on maternal and neonatal health service quality in Malawi. | Non randomized control study. | Health care workers | Malawi | Primary Health care facilities | The scheme improved the availability of both functional equipment and essential drug stocks in the intervention facilities. | 60 |
| 78. | Orubu E.S.F et al; 2019 | EML | Scope availability, price, and affordability of essential cardiovascular medicines for children in selected states in Nigeria. | Longitudinal survey | Health care workers | Nigeria | Hospitals;Pharmacies/Dispensaries | Access to the essential cardiovascular medicines for children were suboptimal in the study locations. | 20 |
| 79. | KakyoT.A et al; 2018 | EML | Understand nurse ward managers perceived challenges in the rural healthcare setting in Uganda. | Qualitative study | Health care managers | Uganda | Hospitals | Health facilities in rural areas face extremely low staff-to-patient ration, a high level of workload, lack of essential medicines and equipment, low salaries and delayed payment for staff. | 60 |
| 80. | Kaiser A.H. et al; 2019 | EML | To explore availability, prices, and affordability of essential medicines for diabetes and hypertension treatment in private pharmacies in three provinces of Zambia. | Cross-sectional study | Health care workers | Zambia | Pharmacies/Dispensaries | The majority of surveyed medicines was inadequately available (<80%),most prices were high in an international comparison and treatment with these medicines was largely unaffordable against the set affordability thresholds. | 80 |
| 81. | Kamuhabwa A et al; 2015 | EML | Assessement of the availability, cost and affordability of essential antibiotics for paediatrics in a semi-rural region in Tanzania. | Cross-sectional study | Health care workers | Tanzania | Hospitals;Primary Health care facilities;Pharmacies/Dispensaries | The median availability of the lowest-price generics of essential antibiotics for paediatrics in the pharmacies, ADDO’s and public hospitals was 59.09%, 62.5%, and 45.5%, respectively | 60 |
| 82. | Marte Y.M. et al; 2018 | EML | Determine the availability and alignment of the Botswana National Essential Medicines List (NEML) for cancer drugs with the WHO’s Essential Medicines List (EML). | Cross-sectional study | Health care workers;Health care managers | Botswana | Hospitals | The 2015 Botswana NEML for cancer had 80.5% alignment with the WHO EML. At least 40% of essential drugs were out of stock for a median duration of 30 days in 2015. Stock outs affected chemotherapy drugs included in first-line regimens for treating potentially curable diseases such as cervical, breast, and colorectal cancer and were not associated with buyer price of therapy. | 60 |
| 83. | Aryeetey G.C. et al; 2016 | EML | Assess the effect of the introduction of the NHIS on health service delivery in mission health facilities in Ghana | Cross-sectional study | Health care managers | Ghana | Hospitals;Primary Health care facilities | All facilities improved the availability of essential medicines after the introduction of the NHIS compared to the period before the NHIS | 100 |
| 84. | Rutebemberwa E. et al; 2016 | EML | To assess the potential of rural and urban private facilities in diagnostic capabilities, operations and human resource in the management of malaria, pneumonia and diarrhoea. | Cross-sectional study | Health care workers | Uganda | Pharmacies/Dispensaries | Rural facilities had less trained personnel and less drug availability. In both rural and urban areas, record keeping was low. | 60 |
| 85. | Wagenaar B.H. et al; 2015 | EML | Assessed the availability of essential medicines for mental healthcare (MH) across levels of the public healthcare system to aid in future systems planning. | Cross-sectional study | Health care workers | Mozambique | Hospitals | Essential psychotropic medicines were often unavailable. No more than half of district warehouses had current availability of any essential typical antipsychotic and no district or individual facilities had availability of the atypical antipsychotic risperidone. | 100 |
| 86. | Maswime T.S. et al; 2016 | EML | To determine preparedness for, and health-system constraints to, safe caesarean section in southern Gauteng hospitals. | Cross-sectional study | Health care workers;Health care managers | South Africa | Hospitals | Services delivery constraints an unequal staff distribution between central hospitals and lower of care, as well as non-availability of essential drugs and a lack of surgical capacity to arrest sever haemorrhage at district and regional hospitals. | 80 |
| 87. | Kumsa A. et al; 2016 | EML | To assess satisfaction with Emergency Obstetric and new born Care services among clients using public health facilities in Jimma zone, Southwest Ethiopia. | Cross-sectional study | Patients | Ethiopia | Primary Health care facilities | The level of clients satisfaction with Emergency Obstetric and New born Care services was low in the study area.  Factors such as availability of essential equipment and drugs, health workers’ communication, health care provided, and attitude of health workers had positive association with client satisfaction. | 100 |
| 88. | Gutema G. et al; 2018 | EML | To determine availability and affordability of commonly prescribed antibiotics at a tertiary hospital in Ethiopia. | Cross-sectional study | Health care workers | Ethiopia | Pharmacies/Dispensaries | Price variations along sector facilities are high as high as over 10 and 19 folds in the private and public pharmacies, respectively, for doxycycline, for instance. | 80 |
| 89. | Demessie M.B. et al; 2020 | EML | This study measured the availability of tracer drugs (TDs) and the implementation of their logistic management information system in public health facilities of Dessie, North-East Ethiopia. | Mixed methods study | Health care workers | Ethiopia | Hospitals;Primary Health care facilities | The average availability of Essential Medicines (EMs) in the public health facilities of Dessie town was 74.7%.  All public health facilities encountered stock out at least by one Tracer Drugs. | 20 |
| 90. | Chukwu.O.A. et al; 2018 | EML | To identify indicators for the poor supply chain management (SCM) for medicines in Nigeria | Mixed methods study | Health care workers;Other | Nigeria | Pharmacies/Dispensaries;Other | Logistics companies are not adequately equipped and no monitoring is in place to ensure that standards are set and followed. | 40 |
| 91. | Okech U.K. et al; 2015 | EML | Evaluate the current status of Intensive Care Unit setups and facilities in Kenya. | Cross-sectional study | Health care workers;Health care managers | Kenya | Hospitals;Primary Health care facilities | Availability and serviceability of ICU equipment, availability of essential drugs and diagnostic support services used in care of critically ill patients ranged from > 95% in private and mission hospitals to 60-80% in the other hospitals | 40 |
| 92. | Treleaven E. et al; 2015 | EML | To assess management of paediatric illnesses by patent and proprietary medicine vendors (PMVs) in Nigeria. | Census | Health care workers | Nigeria | Pharmacies/Dispensaries | Stocking patterns varied by state, urban/rural location and according to whether the ship was headed by someone with formal health training. Many PPMVs lack the knowledge and tools to properly treat common childhood illnesses. | 80 |
| 93. | Armstrong-Hough M et al; 2018 | EML | To model meaningful associations between EM-NCD availability and facility-level characteristics in a sample of Ugandan health facilities. | Cross-sectional study | Health care workers | Uganda | Hospitals | For-profit facilities essential medicines for non-communicable disease counts were 98% higher than public facilities. General hospitals and referral health centers had 98% and 105% higher counts compared to primary health centers. | 80 |
| 94. | Brhlikova P et al; 2020 | EML | To systematically examined the registration and local production of essential medicines | Mixed methods study | Other | Uganda | Others | Local producers faced considerable barriers to achieving international quality standards required for international procurement of medicines for the domestic market. | 80 |
| 95. | Nsabagasani X et al; 2015 | EML | Explore stakeholders’ views about the relevance of the globally recommended child-appropriate dosage formulations in the context of Uganda | Qualitative study | Health care workers;Health care managers | Uganda | Pharmacies/Dispensaries | There has been a slow transition to child-appropriate medicine formulation in Uganda due to resource constraints, lack of sufficient influence of the medicine policy coordinating institutions and a lack of common ground, divergent interests, and values among the stakeholders. | 100 |
| 96. | Perumal-Pillay V.A. et al; 2017 | EML | Aim to refill this gap in knowledge and reported on the decision making process for the selection of essential medicines for SA STG/EMLs and management. | Qualitative study | National Essential List Committees | South Africa | Others | The NEMLC process of medicine selection has been refined over the years. This together with the EML review process is now essentially predominantly an evidence based process where quality, safety and efficacy of a medicine is considered first followed by cost considerations which includes pharmacoeconomic evaluations, and pricing of medicines. | 100 |
| 97. | An.S.J. et al; 2015 | EML | To assess supply-side dimensions and dynamics of integrating HIV testing and counselling into routine antenatal care. | Mixed methods study | Health care workers | Tanzania | Primary Health care facilities | Limitations in structural inputs, such as infrastructure, supplies, and staffing, constrain the potential for integration of HIV testing and counseling into routine antenatal care services. | 40 |
| 98. | Wagenaar B.H. et al; 2014 | EML | To assess the relationship between health system factors and facility-level essential health products (EHPs) stock outs in Mozambique. | Longitudinal analysis | Health care workers | Mozambique | Primary Health care facilities | Facility-level stock-outs of EHPs are common and appear to disproportionately affect those living far from district capital and near facilities with few health staff. | 80 |
| 99. | Berhane A et al; 2015 | EML | To examine patients‚ preference for attributes related to health care services and to ascertain the relative impact of attributes at hospitals in Amhara Region, northern Ethiopia. | Experimental design | Patients | Ethiopia | Hospitals | Participants preferred to visit a hospital which had continuity of care, good physician and nursing communication, full drug availability, and a lot of diagnostic facilities | 100 |
| 100. | Auwal F et al; 2019 | EML | To describe the availability and assess the rationality of selected FDCs available in drug retailing outlets in Kaduna state, Nigeria. | Cross-sectional study | Health care workers | Nigeria | Pharmacies/Dispensaries | There were a wide variety of FDCs available in the study areas. While majority of the FDCs were registered with the Nigerian drug regulatory agency, some of the combinations were found to be irrational. | 60 |
| 101. | Wangu M.M. et al; 2014 | EML | To confirm the level of availability of essential medicines in public hospitals in Kenya and the factors leading to this. | Cross-sectional study | Health care workers | Kenya | Hospitals | Majority of drugs were found to be stocked out. Stock out were caused by low funding of health projects, inappropriate selection and irrational use of drugs. | 60 |
| 102. | Mhlanga B.S. et al; 2014 | EML | To determine the prices, availability and affordability of medicines along the supply chain in Swaziland. | Cross-sectional study | Health care workers | Eswatini (Swaziland) | Pharmacies/Dispensaries | Private sector original brand medicines were priced 32.4 times higher than international reference prices. Standard treatment with originator brands cost more than a day’s wage | 80 |
| 103. | Kiplagat A. et al; 2014 | EML | To identify the factors influencing the implementation of IMCI at Public health centers & dispensaries in Mwanza city. | Cross-sectional study | Health care workers | Tanzania | Primary Health care facilities | The main challenges identified in the implementation of IMCI are low initial training coverage among health care workers, lack of essential drugs and supplies, lack of onsite mentoring and lack of refresher courses and regular supportive supervision. | 80 |
| 104. | Nnebue C.C. et al; 2014 | EML | Determine the adequacy of resources (human and material) for provision of maternal health services at the primary health care (PHC) level in Nnewi, Nigeria | Cross-sectional study | Patients | Nigeria | Primary Health care facilities | The four facilities studied had most of the equipment and drugs available but in insufficient quantities. | 100 |
| 105. | Mujinja P.G. et al; 2014 | EML | To examine the role of locally produced medicines in African markets and on potential benefits of local production for access to medicines | Cross-sectional study | Other | Tanzania | Hospitals;Primary Health care facilities;Pharmacies/Dispensaries | Locally produced medicines are more accessible than imports for rural consumers, and that medicines which are both imported and locally produced display greater rural and urban availability than those which are import-only. | 60 |
| 106. | Birabwa C et al; 2019 | EML | This study assessed the quality of diabetes care in rural health facilities in Uganda | Mixed methods study | Patients;Health care workers | Uganda | Hospitals;Primary Health care facilities | The overall capacity of health facilities to deliver T2DM healthcare was rated above average, while process and outcome performance was poor. | 100 |
| 107. | Koroma M.M et al; 2019 | EML | To evaluate gaps on access to essential medicines and other supplies and equipment necessary for Basic Emergency Obstetrics and Newborn care(BEmONC )services. | Cross-sectional study | Patients;Health care workers;Health care managers | Sierra Leone | Primary Health care facilities | All Peripheral Health Units(PHUs) were available to refer patients for care at a higher-level facility after diagnosing complications during daytime hours. | 60 |
| 108. | Mori A.T et al; 2014 | EML | Describe the process of updating the Standard Treatment Guidelines and National Essential Medicine List in Tanzania and further examines the criteria and the underlying evidence used in decision-making. | Qualitative study | Health care workers;Other | Tanzania | Hospitals;Primary Health care facilities | Efficacy, safety, availability and affordability were the most frequently utilised criteria in decision-making, although these were largely based on experience rather than evidence | 80 |
| 109. | Kayombo E.J et al; 2012 | EML | Aim to get an experience of health care utilization from both urban and rural areas of seven administrative regions in Tanzania. | Qualitative study | Health care managers | Tanzania | Primary Health care facilities | Findings revealed that the health facilities were overburden with higher population to serve than it was planned. | 100 |
| 110. | Magadzire B.P et al; 2014 | EML | Examines in a South African context supply- and demand- ATM barriers from the provider perspective using a five-dimensional framework | Cross-sectional study | Community residents;Health care workers;Health care managers | South Africa | Primary Health care facilities | Availability of medicines was hampered by logistical bottlenecks in the medicines supply chain; poor public transport networks affected accessibility. | 60 |
| 111. | Coghlan R et al; 2018 | EML | Examining the access to essential medicines using the Zambian anti-malarial medicines as a case study. | Cross-sectional study | Other | Zambia | Others | In all years, the supply of products was easily sufficient to meet, and in fact exceeds reported medical need. The public sector did indeed account for the large majority of all imports; up to 98% of value and volume of antimalarials in 2014. | 100 |
| 112. | Rizk H.I et al; 2021 | EML | To identify the determinants of patients‚ access to medicines at the primary health care (PHC) level from the perspectives of various (internal and external) stakeholders of the pharmaceutical system. | Cross-sectional study | Patients;Health care workers | Egypt | Primary Health care facilities | About one-third of patients in both urban and rural facilities were unable to pay for the medicine. The prescribing and dispensing policies are deficiently enforced. | 60 |
| 113. | Obakiro S.B et al; 2021 | EML | To assess affordability and appropriateness of prescriptions written for diabetics patients in Eastern Uganda. | Cross-sectional study | Patients | Uganda | Hospitals | The majority of diabetic patients in Eastern Uganda could not afford to buy prescribed medicines. | 100 |
| 114. | Mbonyinshuti F et al; 2021 | EML | To investigate the availability of essential drugs used in the treatment of selected NCDs | Cross-sectional study | Health care workers | Rwanda | Hospitals;Primary Health care facilities;Pharmacies/Dispensaries | Compared to rural health facilities, NCD drug consumption was higher at the hospital, with no drug stock-outs observed. | 80 |
| 115. | Bouwer J.C et al; 2021 | EML | To describe the profile and cost of medicines procured for managing mental, neurological and substance use (MNS) disorders during the 2017‚2018 financial year. | An exploratory, quantitative analysis | Health care workers | South Africa | Hospitals | Of the total provincial medicines expenditure in 2017–2018, 3.73% was for MNS disorders, which is similar to the spending on cardiovascular (4%) and respiratory (3%) disorders. Antivirals for systemic use comprised 44% of the total expenditure, followed by vaccines at 13% | 60 |
| 116. | Mathewos B et al; 2019 | EML | To assess the strength of the community-based newborn care program implementation in terms of inputs, process, and outputs, and document key lessons learned through the implementation of the first phase of the program in the four agrarian regions of Ethiopia. | Cross-sectional study | Health care workers;Health care managers | Ethiopia | Health care providers | The implementation was in line with the guiding principles and the proposed strategies of the implementation guide.However, were some inconsistencies between what was planned and what actually happened on the ground. | 80 |
| 117. | Dekker AM. et al; 2012 | EML | To examine patient experiences and health care provider attitudes towards chronic pain and palliative care in Eastern Cape Province, South Africa. | Mixed methods study | Patients;Health care workers | South Africa | Hospitals | Factors inhibiting the provision of palliative care included insufficient access and availability of pain medication and providers’ association of palliative care with end-of-life care | 80 |
| 118. | Lukama L et al; 2020 | EML | To investigate ENT service provision in Zambia with regard to availability of surgical procedures and supply of essential drugs. | Cross-sectional study | Health care workers;Health care managers | Zambia | Hospitals | Most hospitals lack essential medication for ENT conditions as listed by the World Health Organization. | 80 |
| 119. | Oduro-Mensah E et al; 2013 | EML | To explore the how‚ and why of care decision making by frontline providers of maternal and newborn services in the Greater Accra region of Ghana and determine appropriate interventions needed to support its quality and related maternal and neonatal outcomes | Cross-sectional study | Health care workers | Ghana | Hospitals;Primary Health care facilities | Health system constraints such as availability of staff, essential medicines, supplies and equipment; management issues (including leadership and interpersonal relations among staff), and barriers to referral were important influences in decision making. | 80 |
| 120. | Obiechina G.O et al; 2013 | EML | To determine students perception of health care services provided in a tertiary institution and assess students‚ attitude towards utilization. | Cross-sectional study | Other | Nigeria | Primary Health care facilities | High cost of drugs, non availability of essential drugs, time spent waiting for treatment, inadequate referral services and satisfaction with services were factors affecting utilization of university health services among students at the university in south‚west Nigeria. | 80 |
| 121. | Hailu A.D. et al; 2020 | EML | To assess the availability, price, and affordability of WHO priority maternal and child medicines in public health facilities, Dessie, North-East Ethiopia. | Cross-sectional study | Health care workers | Ethiopia | Hospitals;Primary Health care facilities | The average mean availability of WHO prioritized MCH medicines in the past 6 months and point mean availability was very low and a high number of stock-outs. | 100 |
| 122. | Awucha N.E et al; 2020 | EML | To examine the impacts of the COVID-19 pandemic on the ease of access to essential medicines by end users. | Cross-sectional study | Patients | Nigeria | Patients | There were difficulties in accessing essential medicines for chronic illnesses with a resultant worsening of chronic conditions, increased medicine costs due to supply chain disruptions, decrease in income generation to make out-of-pocket payments for medicines, and a shift from where medicines were sourced. | 60 |
| 123. | Ashigbie P.G et al; 2020 | EML | To determine the availability and prices of medicines for non-communicable diseases (NCDs) in health facilities and private for-profit drug outlets in Kenya. | Cross-sectional study | Health care workers | Kenya | Hospitals;Primary Health care facilities;Pharmacies/Dispensaries | The availability of NCD medicines in Kenya is significantly lower than the target level of 80%. Availability is poorest in the public sector, and generally highest in the private for-profit sector. | 60 |
| 124. | Seid S.S et al; 2018 | EML | To identify the factors compelling the execution of integrated management of neonatal and childhood illness (IMNCI) by nurse in four districts of West Arsi zone of Ethiopia. | Cross-sectional study | Health care workers  \ | Ethiopia | Hospitals;Primary Health care facilities | The most common issues encountered in the implementation of IMCI were: lack of trained staff (56.2%), lack of essential drugs and supplies (37.3%), and irregular supportive supervision (89.2%) | 60 |
| 125. | Miltenburg A.S et al; 2017 | EML | To perform a district-wide assessment of emergency obstetric and newborn care performance and identify ways for improvement. | Cross-sectional study | Health care workers | Tanzania | Hospitals;Primary Health care facilities;Pharmacies/Dispensaries | Inadequate availability of essential drugs, lack of ability to perform vacuum extraction and blood transfusion limited the performance of emergency obstetric and newborn care. | 40 |
| 126. | Perumal-Pillay V.A et al; 2017 | EML | To gather opinions and perceptions from parents/guardians on availability, affordability and quality of medicines and healthcare for children in SA. | Qualitative study | Community residents | South Africa | Community residents | Medicines and healthcare facilities are accessible in urban and peri-urban areas. medicines may not always be available in public sector facilities due to medicine shortages, compelling parents to purchase medicines from private sector pharmacies. | 100 |
| 127. | Oyekale A.S et al; 2017 | EML | To analyze service readiness of Primary Health Care (PHC) facilities in Nigeria with focus on availability of some essential drugs and medical equipment. | Cross-sectional study | Health care workers | Nigeria | Hospitals;Primary Health care facilities;Pharmacies/Dispensaries | There is some variances between availability of basic medical equipment and their functionality. It was also noted that basic drugs are not readily available at the selected health facilities. | 80 |
| 128. | Namuyinga R.J et al; 2017 | EML | To determine appropriate malaria testing and treatment practices four years after implementation of a policy requiring diagnostic confirmation before treatment. | Cross-sectional study | Patients;Health care workers | Malawi | Hospitals;Community services | The majority of the health facilities had at least on AL formulation available (Ninety-six (91%) HFs had either the first-line (AL) or second-line (artesunate-amodiaquine (ASAQ)) ACT available for the full day of the survey). | 100 |
| 129. | Ooms G.I et al; 2019 | EML | To identify the social, cultural, and regulatory barriers that influence access to internationally controlled essential medicines in Uganda. | Qualitative study | Health care managers | Uganda | Hospitals;Other | ICEM-specific barriers in Uganda were due to nonprioritization of ICEMs, difficulties in finding a balance between control and access, lack of knowledge among health care providers and the population, and stigma. | 100 |
| 130. | Olubumni O.M et al; 2019 | EML | To determine the perceptions of professional nurses regarding the status of stock-outs of generic medicines at primary health care health facilities in a selected province of South Africa. | Qualitative study | Health care workers | South Africa | Primary Health care facilities | Key findings showed that essential medicines were not always available, with the health centres reporting fewer stock-outs than clinics. The perceived major contributors to stock-outs were institutional inefficiency and practices by both health service providers and patients. | 100 |
| 131. | Maïga D et al; 2010 | EML | To assess the impact of this intervention on the evolution of market prices and the subsequent availability and public access to essential medicines in Mali. | Cross-sectional study | Health care workers | Mali | Pharmacies/Dispensaries;Other | There was a small degree of variation in the wholesale and retail prices; median wholesale prices were 25.6% cheaper than those fixed by the regulation, while median retail prices were only 3% more expensive than the decreed maximum prices. | 60 |
| 132. | Khuluza F et al; 2019 | EML | To investigate the prices,affordability and availability of essential medicines in Malawi. | Cross-sectional study | Health care workers | Malawi | Hospitals;Primary Health care facilities;Pharmacies/Dispensaries | Low availability of essential medicines in the public sector especially for pediatric dosage formulations and for a range of NCD medications. | 100 |
| 133. | Daniel G. et al; 2011 | EML | To obtain preliminary data on the drug supply management system in Ethiopia. | Mixed methods study | Health care workers | Ethiopia | Primary Health care facilities | Inadequate availability of essential drugs and commodities in the surveyed facilities as well as weakness in human resources and training. | 80 |
| 134. | Kefale A.T et al; 2019 | EML | To assess essential medicines (EMS) availability and inventory management practices at health centers (HCs) of Adama town, Ethiopia. | Cross-sectional study | Health care workers | Ethiopia | Primary Health care facilities | The availability of EMs and the accuracy of record keeping in the HCs were low. The major problem common for all HCs in the procurement process were PFSA stock status and transportation. | 80 |
| 135. | Epiu I et al; 2017 | EML | To identify key bottlenecks in the provision of safe obstetric anaesthesia | Cross-sectional study | Health care workers | Uganda | Hospitals;Primary Health care facilities | Shortage of anaesthetic supplies were reported throughout the country, and other essential drugs such as anti-acids, anti-hypertensive drugs, opioids and non-opioids, etc. | 80 |
| 136. | Chukwu O.A et al; 2016 | EML | To assess the awareness and readiness of Nigerian pharmacists on supply chain management practices for improving access to medicines. | Cross-sectional study | Health care workers | Nigeria | Hospitals;Pharmacies/Dispensaries | Pharmacists in Nigeria have limited awareness and readiness on supply chain management of health commodities in Nigeria. | 60 |
| 137. | Sado E et al; 2016 | EML | To assess access to essential medicines (EMs) for children based on availability, affordability, and price. | Cross-sectional study | Health care workers | Ethiopia | Pharmacies/Dispensaries | Access to EMs to children is hampered by low availability and high price. Medicines were unaffordable for treatment of common conditions prevalent in the zone at both public and private sectors. | 100 |
| 138. | Mackintosh M et al; 2010 | EML | Evaluation of Markets and Policy Challenges in Access to Essential Medicines for Endemic Disease | Mixed methods study | Other | Tanzania | Hospitals;Primary Health care facilities;Pharmacies/Dispensaries | Wholesale market competition, in contrast, should be promoted, while the rebuilding of African pharmaceutical manufacturing is important for promoting and sustaining access. | 100 |
| 139. | Gilroy K.E et al; 2012 | EML | Assess the quality of care provided by Health Surveillance Assistants (HSAs) cadre of community-based health workers’s as part of a national scale-up of community case management of childhood illness (CCM) in Malawi. | Cross-sectional study | Health care workers;Other | Malawi | Health care workers | HSAs correctly assessed 37% of children for four danger signs by conducting a physical exam, and correctly referred 55% of children with danger signs. | 80 |
| 140. | Sambo L.G. et al; 2011 | EML | To reflect on the discussions held during the Fifteenth Assembly of the Heads of State and Government of the African Union on the reasons why the maternal mortality ratio is so high in Africa and what can be done to reduce it. | Panel and Open Public discussions | Health care stakeholders | Uganda | Health care stakeholders | There is high maternal mortality ratios in countries were attributed to weak national health information systems, leadership and governance challenges | N/A |
| 141. | Modisakeng C et al; 2020 | EML | Highlight challenges in the current pharmaceutical procurement process for public sector hospitals. | Qualitative study | Health care workers;Health care managers;Other | South Africa | Hospitals;Primary Health care facilities;Pharmacies/Dispensaries | The challenges experienced with medicines shortages did not discriminate against the different categories of the hospitals as they were similar across all levels of care. | 100 |
| 142. | Khuluza F et al; 2017 | EML | Analysis of availability and prices of antimalarial and antibiotic medicines in public, faith-based and private health facilities in Malawi. | Cross-sectional study | Health care workers | Malawi | Hospitals;Primary Health care facilities;Pharmacies/Dispensaries | Availability of the antimalarial which are provided with financial support from international donors, was high in public and CHAM facilities. | 80 |
| 143. | Lyon C.B et al; 2016 | EML | To evaluate the anaesthesia capacity of Mozambique | Cross-sectional study | Health care workers;Health care managers | Mozambique | Hospitals | There is minimal capacity for growth given only 1 Mozambique anesthesia residency with inadequate resources. The most commonly perceived barriers to safe anesthesia in this critical shortage are lack of teachers, lack of medical student interest in and exposure to anesthesia, need for more schools, low allocation to anesthesia from the list of available specialist prospects by MOH, and low public payments to anesthesiologists. | 60 |
| 144. | Kusemererwa D et al; 2016 | EML | To explore the relationships among government funding allocations for EMHS, patient load, and medicines availability across facilities at different levels of care. | Cross-sectional study | Health care workers | Uganda | Hospitals;Primary Health care facilities;Community services | There were wide variations in EMHS allocations per patient within the same levels of care. HC IVs and regional referral hospitals had the widest disparities in patient load and therefore in allocation per patient. | 100 |
| 145. | Muhammed K.A et al; 2013 | EML | Identify the predominant barriers affecting the utilization of primary health care services in Batsari Local Government in Katsina State, Nigeria | Cross-sectional study | Community residents | Nigeria | Primary Health care facilities | Some of the multiple factors affecting the utilization of primary healthcare services included lack of essential drugs, high cost of services as well as inadequate infrastructure. | 80 |
| 146. | Lonnée HA  et al; 2017 | EML | To assess the type of anesthesia used for cesarean delivery, the level of training of anesthesia providers, and to document the availability of essential anesthetic drugs and equipment in provincial, district, and mission hospitals in Zimbabwe | Cross-sectional study | Health care workers | Zimbabwe | Hospitals | Provincial hospitals, where district/mission hospitals refer difficult cases, did not have the higher level anesthesia expertise required to manage these cases. | 100 |
| 147. | Abiye Z et al; 2013 | EML | To find out the availability and price of selected medicines as well as affordability of cost of treatment for common diseases in Jimma Health Center (JHC) pharmacy. | Cross-sectional study | Patients | Ethiopia | Primary Health care facilities | Overall availability of medicines in JHC was very low. Consequently, patients were forced to purchase drugs at higher prices in private pharmacies or go to informal sector or forgo treatment. | 80 |
| 148. | Callaghan-Koru J.A et al; 2013 | EML | To assess how health systems are supporting the national community-based health worker (CBHW) to improve access to primary health care. | Mixed methods study | Health care workers;Health care managers;Other | Malawi | Primary Health care facilities | Among the 131 HSAs enrolled in the survey, 68.7% had all three critical drugs available at their respective community case management (CCM) clinics at the time of the survey team visit. | 80 |
| 149. | Amadi C. et al; 2018 | EML | To examine stakeholder perspectives on quality control of essential medicines for maternal and child health, while characterizing pharmaceutical distribution of medicines in Nigeria. | Mixed methods study | Health care workers | Nigeria | Primary Health care facilities | Identified challenges in assuring quality included existence of open market and failed policy implementation, illicit sale of prescription medicine by patent medicine vendors, insufficient regulatory agencies. | 100 |
| 150. | Bizimana T et al; 2020 | EML | Provide data on prices, availability and affordability of medicines in different health facilities of Rwanda | Cross-sectional study | Health care workers | Rwanda | Hospitals;Primary Health care facilities;Pharmacies/Dispensaries | Prices for generic medicines in public and faith-based health facilities were remarkably low, with median price ratios (MPRs) of 1.0 in comparison to the international procurement prices. | 100 |
| 151. | Yevutsey S.K et al; 2017 | EML | Situational analysis of antimicrobial use and resistance in Ghana, with focus on policy and regulation | Situational analysis | Other | Ghana | Hospitals | There was no National Antimicrobial Policy (NAP). Antibiotics are widely prescribed and dispensed by unauthorized persons, suggesting weak enforcement of the law. | 100 |
| 152. | Karanja S et al; 2018 | EML | To determine factors influencing health facility delivery and describe barriers and motivators to the same | Mixed methods | Patients;Health care workers;Other | Kenya | Health care stakeholders | High parity was negatively associated with health facility delivery. Barriers to health facility delivery included women not being final decision makers on place of birth, lack of a birth plan, gender of health provider, unfamiliar birthing position, disrespect and/or abuse, distance, attitude of health providers and lack of essential drugs and supplies. | 60 |
| 153. | Visser C.A et al; 2018 | EML | Evaluate the quality of care provided, the barriers to the effective rollout of antiretroviral services and the role of a clinical mentor. | Mixed methods study | Patients;Health care workers | South Africa | Primary Health care facilities | The quality of care provided was comparable to doctor-monitored care. The facility audits found recurrent shortages of essential drugs. Patients indicated a high level of satisfaction. Salary challenges,excessive workload, a lack of trained nurses and infrastructural barriers were identified as barriers | 100 |
| 154. | Patel A et al; 2012 | EML | To compare South African consumers‚ and healthcare providers‚perceptions of quality of generics to the actual quality of selected products. | Qualitative study | Health care workers;Other | South Africa | Pharmacies/Dispensaries;Other | Procurement and use behavior of healthcare providers was influenced by prior experience, manufacturers’ names and consumers’ ability to pay. All formulations passed the in vitro tests for quality. | 100 |
| 155. | Birabwa C. et al; 2014 | EML | This study looks at the availability of six essential medicines in private drug outlets across Uganda. | Cross-sectional study | Health care workers | Uganda | Pharmacies/Dispensaries | This study shows that some essential medicines are quite common in private outlets and sold in drug shops that do not have a license for sale. | 80 |
| 156. | Robertson J et al; 2015 | EML | To examine the consistency of the results across the studies and to compare the resulting inferences on access to essential medicines for these NCDs | Cross-sectional study. | Health care workers | Tanzania | Hospitals;Primary Health care facilities;Pharmacies/Dispensaries | The availability of key NCD medicines for the management of diabetes and hypertension is suboptimal. There were consistent patterns of lower availability in government facilities than mission/faith-based and private facilities | 80 |
| 157. | Exley J. et al; 2016 | EML | To understand, from multiple perspectives, the views and experiences of childbearing women living in areas where it has been implemented. | Qualitative study | Community residents;Health care workers;Other | Nigeria | Hospitals;Primary Health care facilities | There had been positive improvements in maternity care. However, despite improvements in the perceived quality of care and an apparent willingness to give birth in a PHC, more women gave birth at home than intended. | 80 |
| 158. | Crowley T et al; 2015 | EML | To evaluate the adequacy of pharmaceutical services for the provision of ART in PHC clinics. | Cross-sectional study | Health care workers | South Africa | Primary Health care facilities | The pharmaceutical services in all the assessed PHC clinics were inadequate according to the basic HIV standards for PHC facilities.None of the clinics complied with all the standard criteria. | 80 |
| 159. | Jingi A.M et al; 2014 | EML | To assess the availability and affordability of medicines and routine tests for cardiovascular disease (CVD) and diabetes in the West region of Cameroon, a low-income setting. | Cross-sectional study | Health care workers;Other | Cameroon | Hospitals;Pharmacies/Dispensaries | The investigation and management of patients with medium to-high cardiovascular risk remains largely unavailable and unaffordable in this setting. | 60 |
| 160. | Oyinlade O.A et al; 2014 | EML | To evaluate school health services in public and private schools in Sagamu, Nigeria | Cross-sectional study | Other | Nigeria | Pharmacies/Dispensaries | All the public primary schools had first aid boxes and essential drugs and materials. Among 50 (94.3%) private schools which had first aid boxes, 47 (88.7%) had essential drugs and materials | 60 |
| 161. | Ozoh B.O. et al; 2020 | EML | To determine the availability and affordability of asthma and COPD medicines across Nigeria. | Cross-sectional study | Health care workers | Nigeria | Hospitals;Pharmacies/Dispensaries | The available medicines were oral corticosteroids and oral salbutamol which are not on the WHO Essential Medicine List. None of the EML medicines were affordable. | 80 |
| 162. | Hamel M.J et al; 2011 | EML | To explore factors that may have contributed to changes in mortality. | Longitudinal analysis | Patients;Community residents;Health care workers | Kenya | Hospitals | Mortality remained elevated during the first 8 months of 2009 compared with 2006 and 2007. Malaria and/or anemia accounted for the greatest increases in child mortality. Stock-outs of essential antimalarial drugs during a time of increased malaria transmission and disruption of services during civil unrest may have contributed to increased mortality in 2008–2009. | 60 |
| 163. | Onyango M.A. et al; 2018 | EML | To understand patients perceptions of access to medicines; as well as barriers and facilitators at the household, community, and healthcare system level. | Qualitative study | Patients | Kenya | Primary Health care facilities | Although medicines at government facilities were free or cheaper than those sold in private pharmacies, the availability of medicines presented a constant challenge. Patients often resorted to private pharmacies, where NCD medicines cost more than at public facilities. | 100 |
| 164. | Mkoka D.A et al; 2014 | EML | To describe the experience of rural health facility managers in ensuring the timely availability of drugs and medical supplies for emergency obstetric care (EmOC). | Qualitative study | Health care workers;Other | Tanzania | Primary Health care facilities;Pharmacies/Dispensaries | The unreliability of obtaining drugs and medical supplies was reported to result in the provision of untimely and suboptimal EmOC services. An insufficient budget for drugs from central government, lack of accountability within the supply system and a bureaucratic process of accessing the locally mobilized drug fund were reported to contribute to the current situation. | 100 |
| 165. | Saouadogo H et al; 2010 | EML | To assess the availability of the essential medicines and the hospital pharmacy management quality | Cross-sectional study | Patients;Health care workers;Health care managers | Burkina Faso | Hospitals | The essential medicines availability and affordability are low. The essential medicines are more available and affordable in the CHRs than the CHUs. | 20 |
| 166. | Boachie M 2015 | EML | To investigate the factors that enrolees in the Ashanti Region considered in choosing preferred primary healthcare providers (PPPs) and direction of association of such factors with the choice of PPP. | Cross-sectional study | Patients | Ghana | Primary healthcare providers | Shorter waiting time, availability of essential drugs and doctors, proximity, recommendations, clean environment and presence of additional charges were the criteria adopted by enrolees in choosing their PPPs. | 80 |
| 167. | Nsabagasani X. et al; 2016 | EML | To explore healthworkers‚knowledge and perspectives about child-appropriate dosage formulations, their practices, and experiences in the management of childhood illnesses in Uganda using IMCI approach. | Qualitative study | Health care workers | Uganda | Hospitals | Child-appropriate dosage formulations were not included in the package for the Integrated Management of Childhood Illnesses and ministry officials attributed this to resource constraints and lack of initial guidance from the World Health Organization. | 100 |
| 168. | Loveday M et al; 2011 | EML | To report on a participatory quality improvement intervention designed to evaluate these priority programmes in primary health care (PHC) clinics in a rural district in KwaZulu-Natal. | Cross-sectional study | Health care workers | South Africa | Primary Health care facilities | Aspects of availability, capacity and continuity of care as hindering effective service delivery: key drugs, stocks and equipment were not available; capacity of staff was compromised by inadequate training and supervision; and continuity of care was hindered by poor laboratory support and no recall system. | 100 |
| 169. | Mubyazi G.M . et al; 2013 | EML | To elucidates the perspectives of District Council Health Management Team(CHMT)s regarding the feasibility of IPTp with SP strategy. | Cross-sectional study | Patients | Tanzania | Primary Health care facilities | The identified weaknesses include late arrival of funds from central level weakening CHMTs performance in health supervision, organizing outreach clinics, distributing essential supplies, and delivery of IPTp services. | 100 |
| 170. | Saksena P et al; 2010 | EML | To identify the magnitude of out-of-pocket payments incurred by caretakers and arising from paediatric inpatient episodes in relation to family income | Cross-sectional study | Patients | Tanzania | Hospitals | The financial cost of hospitalization for under-fives can be a considerable burden on households in Tanzania, with a mean payment of US$5.5. | 40 |
| 171. | Nakyanzi J.K et al; 2010 | EML | To investigate the extent of, and the main contributing factors to, expiry of medicines in medicine supply outlets in Kampala and Entebbe, Uganda. | Cross-sectional study | Health care workers | Uganda | Pharmacies/Dispensaries | Expiry of medicines in supply facilities was common among medicines for vertical health programmes. Poor coordination appears to be responsible for some expiry incidents | 80 |
| 172. | Nilseng J et al; 2014 | EML | Assess current approaches to and use of ICT among health workers in two rural districts of Tanzania in relation to the current drug distribution practices, drug stock and continuing medical information (CME), as well as assessing the feasibility of using ICT to improve ordering and use of medicines. | Qualitative study | Health care workers | Tanzania | Hospitals;Primary Health care facilities;Pharmacies/Dispensaries | The Health workers were very positive to the tablet application and saw its potential in reducing drug stock-outs. They also expressed a great need and wish for CME by distance. | 100 |
